# Spatially resolved proteomic signatures of atherosclerotic carotid artery disease

**DOI:** 10.1101/2025.02.09.25321955

**Authors:** Ankit Sinha, Nadja Sachs, Elena Kratz, Jessica Pauli, Sophia Steigerwald, Vincent Albrecht, Thierry Nordmann, Enes Ugur, Edwin H Rodriguez, Marie-Luise Engl, Patricia Skowronek, Moritz von Scheidt, Hanna Winter, Daniela Branzan, Heribert Schunkert, Lars Maegdefessel, Matthias Mann

## Abstract

Atherosclerotic plaque rupture remains a leading cause of adverse cardiovascular events, yet the molecular drivers of plaque vulnerability are incompletely understood. To address this challenge, we developed an integrated approach that combines histomorphology-guided spatial proteomics with machine learning to map protein signatures across spatially distinct plaque subregions. Our analysis revealed that vulnerability signatures concentrate in the necrotic core and fibrous cap subregions, and are significantly enriched for ossification, inflammation, cholesterol metabolism, and extracellular matrix degradation pathways. When comparing the vulnerability status across subregions, we found that the necrotic core has the most distinctive vulnerability-associated proteome, with 454 proteins significantly altered between stable and vulnerable states. We identified a mechanistic link between inflammation and oxidative stress, PCSK9 upregulation, and vascular smooth muscle cell dysfunction in vulnerable plaques. This finding suggests arterial PCSK9 as a therapeutic target beyond its established role in hepatic lipid metabolism. By employing machine learning, we developed and independently validated a seven-protein tissue panel (receiver operating characteristic– area under the curve = 0.86) and found a 12-protein serum panel to predict plaque vulnerability status. Thus, plaque vulnerability signatures are spatially concentrated in specific subregions and highlight actionable biomarkers and therapeutic targets for advanced carotid artery disease.

## Introduction

The rupture of vulnerable carotid plaques often leads to devastating cerebrovascular events. Detecting these high-risk lesions remains a valuable yet elusive clinical challenge. Although severe arterial stenosis (>75% luminal occlusion) was once the primary indicator for stroke risk and intervention decisions, recent evidence shows that a large proportion of acute events are driven by plaques with moderate stenosis (50– 70% occlusion)^1–3^. This shift from flow restriction to a plaque composition paradigm underscores that plaques with varying degrees of stenosis can harbour morphological features (e.g., intraplaque haemorrhage, large lipid necrotic cores, thin fibrous caps) and molecular drivers that predispose them to rupture^4^. This realisation highlights the importance of molecular profiling (e.g., transcriptomic and proteomic) for elucidating the cellular and extracellular mechanisms underlying plaque vulnerability^5,6^. Thus, a deeper understanding of molecular composition, along with spatial heterogeneity, may further uncover new mechanisms of plaque vulnerability and provide the needed reliable biomarkers for risk stratification and precise intervention.

Carotid plaques are not spatially homogeneous; rather, they comprise three key subregions—the tunica media, a smooth muscle–rich layer providing tensile strength; the necrotic core, a lipid-rich region replete with apoptotic cells and debris; and the fibrous cap, a collagenous barrier that separates the thrombogenic necrotic core from the bloodstream^7–9^. The distinct cellular and extracellular composition of these subregions likely influences plaque stability. For example, fibrillar collagens (types I and III) secreted by synthetic vascular smooth muscle cells (VSMCs) strengthen the fibrous cap, whereas the necrotic core harbours pro-inflammatory proteins (e.g., TNFα, IL-1β, MMPs) and infiltrating immune cells (e.g., macrophages, T-cells) that exacerbate plaque inflammation and matrix degradation^10,11^. Meanwhile, the media harbours contractile VSMCs and a highly structured extracellular matrix (ECM; e.g., elastin, collagens) essential for maintaining arterial wall integrity^12^. These distinct molecular and cellular signatures across plaque subregions underscore the need for spatial molecular analyses to elucidate how their interactions influence plaque stability and rupture risk.

Although prior transcriptomic and proteomic studies have correlated molecular alterations in carotid artery disease, bulk analyses do not capture the localised molecular changes within plaque subregions^13–15^. Consequently, the biological contributions of each subregion to overall plaque vulnerability remain unclear^16^. To bridge this gap, we applied mass spectrometry (MS)–based spatial proteomics to investigate protein changes associated with plaque vulnerability across the three subregions. A spatial proteomics approach can reveal localised cellular and extracellular changes, thereby improving the detection of molecular drivers of plaque vulnerability by increasing the effective resolution of proteomic data.

We hypothesise that distinct protein signatures within plaque subregions orchestrate vulnerability through spatially organised biological programmes. We systematically investigate how (1) subregions are differentially associated with plaque vulnerability status, (2) the biological processes drive subtype heterogeneity within each subregion, and (3) the aggregate of subregion and subtype information provides robust protein signatures for predicting vulnerability status. By coupling histomorphological spatial context with high-resolution quantitative proteomics, this study reveals novel molecular determinants of plaque vulnerability and establishes a framework for developing spatially informed protein signatures.

## Results

### Study design and clinical cohorts

To identify protein signatures associated with atherosclerotic plaque vulnerability, we established a histomorphology-guided proteomics workflow (Fig. 1A). Guided by a pathologist, we used laser microdissection to isolate the spatially distinct plaque subregions—media, necrotic core, and fibrous cap—from carotid plaques of patients undergoing carotid endarterectomy (CEA). We profiled the proteomes of each subregion and patient using a customised acquisition method for ion-mobility mass spectrometry^17^. This workflow enabled precise sampling while preserving the correlation between protein abundance and spatial subregions in stable and vulnerable plaques.

**Figure 1.**
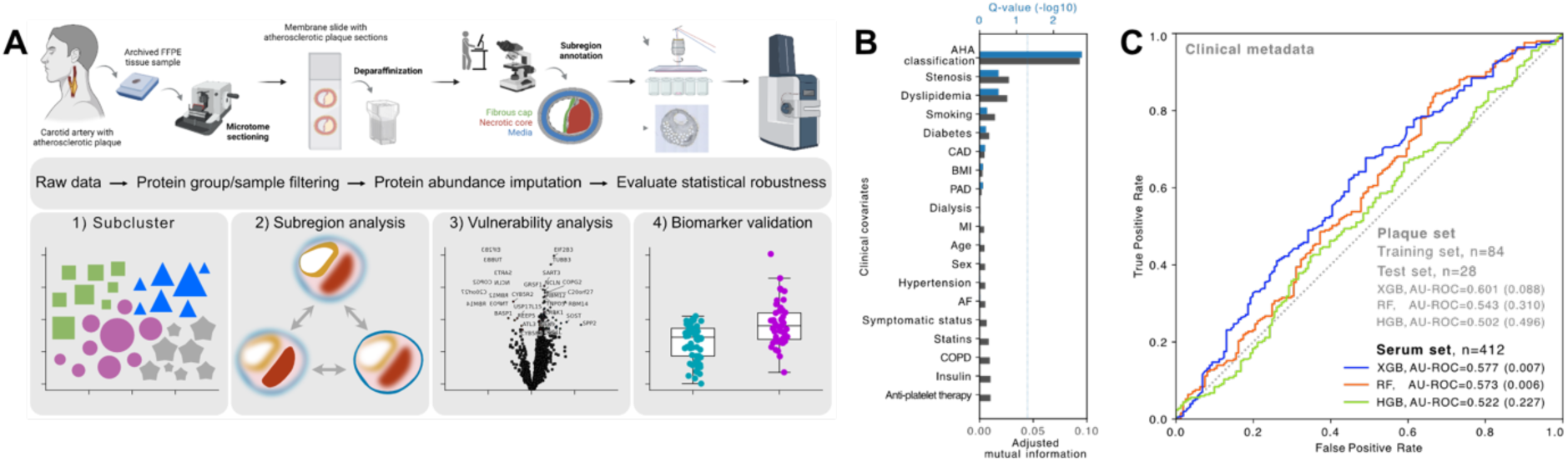
Proteomic phenotyping of carotid plaque instability reveals distinct molecular phenotypes. **A**, Workflow for proteomic analysis of carotid endarterectomy specimens (n=12), including microdissection of histologically defined plaque regions, quality-controlled protein quantification, and integrated data analysis. **B**, Clinical covariates ranked by their association with plaque vulnerability, quantified by normalised mutual information (black bars). Blue bars indicate statistical significance (Q < 0.05, dotted line). **C**, Assessment of plaque vulnerability prediction using clinical data alone. Three machine learning models (XGBoost, Random Forest, Histogram Gradient Boosting) showed limited predictive power (AU-ROC: 0.50-0.60, p value determined using label permutation in parentheses), highlighting the need for molecular markers. Figure created with BioRender.com.

We applied this workflow to a cohort of carotid plaques from 112 patients. Cases were selected based on the completeness of clinical data, clinical covariate class balance, and the histological quality of the media subregion. Clinical covariates were matched to ensure comparability between stable (n=55) and vulnerable (n=57) plaque. The vulnerable status was defined by fibrous cap thickness <200 μm^18^. Other covariates included dyslipidaemia, diabetes, BMI, sex, symptom status (asymptomatic vs symptomatic for transient ischemic attack, amaurosis fugax or stroke), medication, and lifestyle history (Table 1). Notably, 94% of cases had an American Heart Association (AHA) histological classification^19^ of V or higher, and 79% had severe stenosis (≥80% luminal occlusion). In our clinical metadata, AHA classification was the only covariate correlated with vulnerability status, albeit weakly (adjusted mutual information, AMI = 0.095, P = 0.003, permutation test; Fig. 1B). Furthermore, clinical covariates, either individually or in combination, had limited accuracy in predicting plaque vulnerability status (receiver operating characteristic–area under the curve, AUC= 0.50–0.60, Fig. 1C). We further verified the limited predictive performance of the clinical covariates in a larger “serum-only” cohort (n=412, AUC = 0.52–0.58, Fig. 1C). Therefore, the poor predictive performance of clinical metadata underscores the need for protein-based signatures of plaque vulnerability.

**Table 1.**
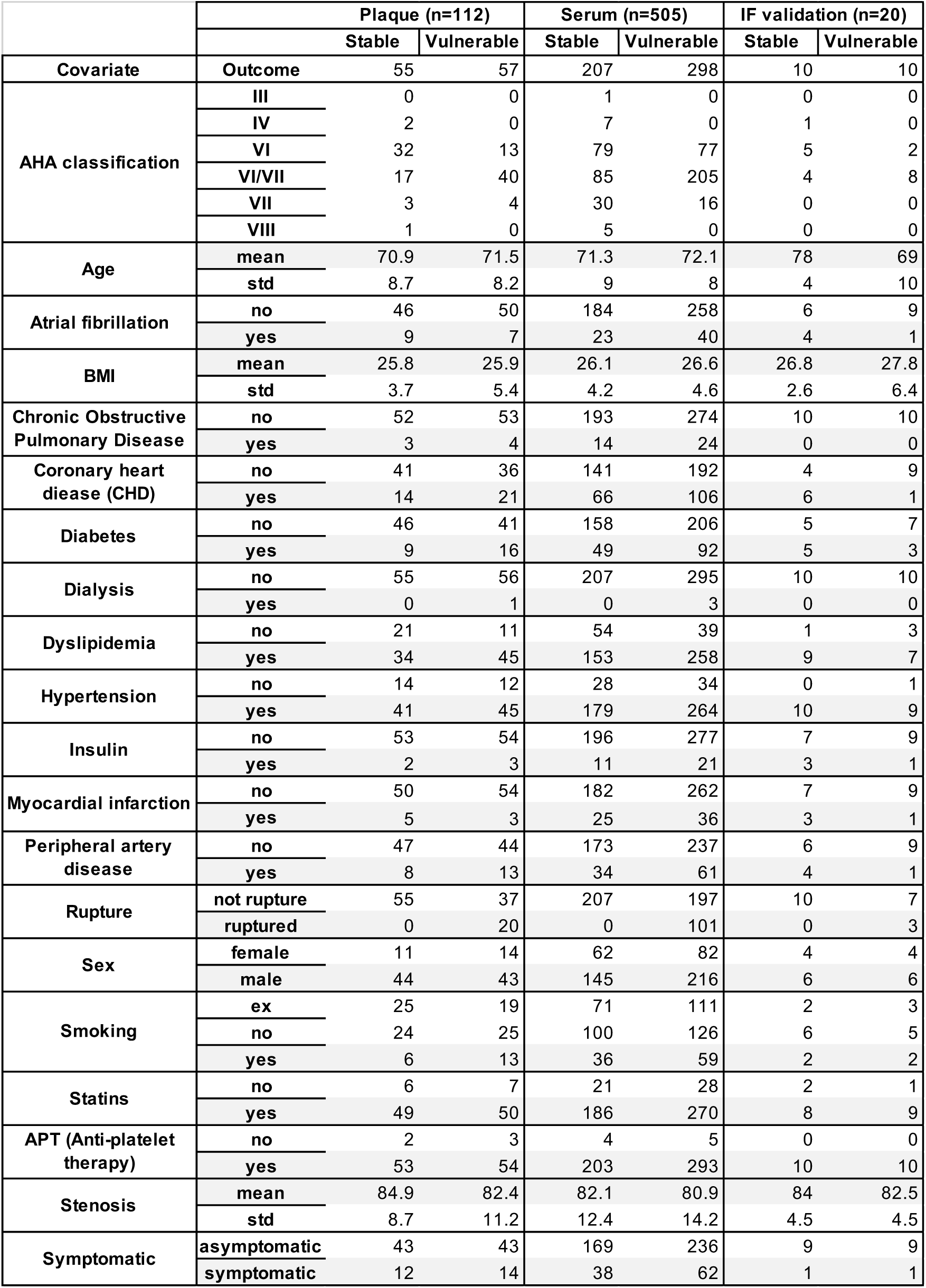
Overview of the clinical covariates for the three patient cohorts used in the study.

### Quantitative patterns in plaque proteomes

Our proteomic analysis identified 4,893 unique proteins across all subregions. The subregion-specific filtering based on observed quantitation yielded an average of 3,412 proteins in the media (n=106), 3,321 in the necrotic core (n=110), and 2,957 in the fibrous cap (n=102) from stable and vulnerable plaques (Fig. 2A). To create a complete data matrix for integrative analyses, we refined the protein quantitation process to minimise statistical artefacts in two ways. Firstly, we restricted quantitation to a set of highly correlated peptides for each protein using internal consistency analysis (Extended Fig. 1A, B). When tested in the proteomics data of a cellular model with known treatment and control conditions, our approach increased effect sizes for the significantly altered proteins (Extended Fig. 1C, D). Secondly, to create a finalised matrix, we used a random-forest-based chained equations technique to impute missing values as it had the smallest imputation artefact (Extended Fig. 1E)^20^.

**Figure 2.**
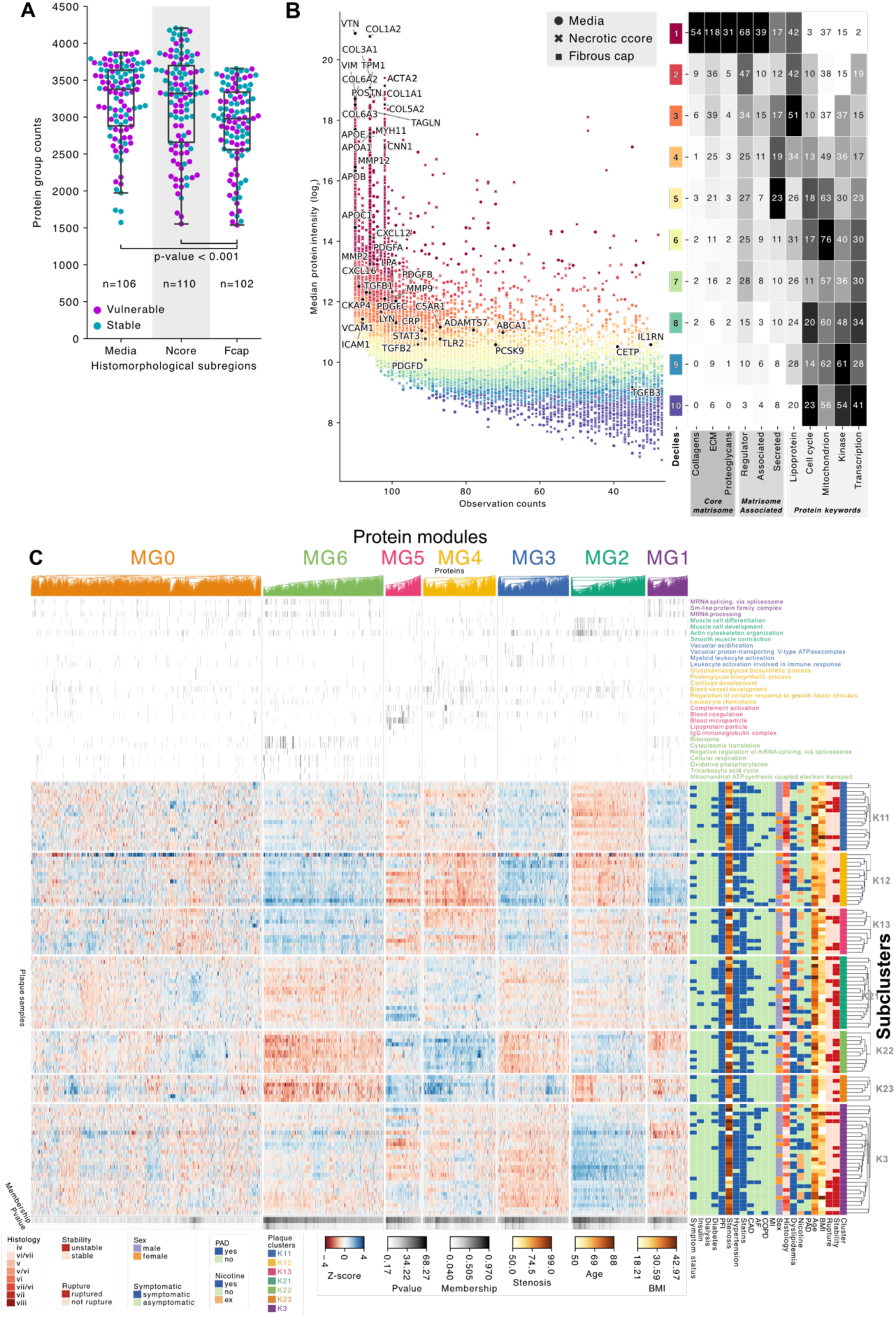
Proteomic phenotyping of carotid plaque instability reveals distinct molecular phenotypes. **A**, Distribution of quantified proteins across plaque subregions after quality filtering. Proteins with more than 66% missing values, and samples with fewer than 1500 proteins were excluded. **B**, Protein abundance patterns across plaque subregions. Colour scale represents protein abundance deciles; right panel displays protein class distributions across deciles. **C**, Consensus clustering analysis of necrotic core samples (n=110) using the top 66% most variable proteins, yielding seven subclusters (K=7). Clinical covariates are shown in row annotations (right). Weighted gene correlation network analysis identified seven protein modules (molecular groups, MG0-MG6) and are shown as top column annotation. MG0 contains low membership (unassigned) proteins. Bottom column annotations indicate protein module membership and the statistical significance of the membership.

To investigate how protein abundance distributions relate to biological processes in advanced (stable and vulnerable) plaques, we stratified proteins by abundance deciles (based on MS-derived median intensity) and functional categories (UniProt keywords). This high-resolution analysis revealed distinct abundance compartmentalisation of the plaque proteome, with structural proteins occupying the highest decile (decile 1; Fig. 2B, right). These included ECM components (e.g., VTN, COL1A2, COL3A1, COL6A2) and VSMC contractility markers (e.g., TPM1, ACTA2, TAGLN, MYH11; Fig. 2B, left). Proteins involved in ECM remodelling and inflammatory or immune signalling pathways were most prevalent in the intermediate deciles (deciles 2–7), whereas intracellular regulatory proteins (e.g., cell cycle modulators, kinases) occupied the lower deciles (deciles 6–10)^21^. These patterns underscore a fundamental connection between protein abundance and plaque homeostasis, wherein the coordinated expression of structural and regulatory proteins governs plaque lesion stability and tissue adaptation.

Having established that protein abundance patterns align with functional groups, we next examined whether plaque subregions have localised protein abundance patterns that correlate with specific clinical covariates or biological processes. We detected seven subclusters in the necrotic core using unsupervised, recursive consensus clustering of plaques (K11–K13, K21–K23, and K3; Fig. 2C, Extended Fig. 2A). Although body mass index (adjusted mutual information, AMI = 0.03, *P* = 0.10), age (AMI = 0.025, *P* = 0.14), and vulnerability status (AMI = 0.021, *P* = 0.08) showed the highest associations with these subclusters, none reached statistical significance (Extended Fig. 3A). In contrast, fibrous cap subclusters (K = 6, Extended Fig. 2B) were significantly associated with chronic obstructive pulmonary disease (COPD; AMI = 0.032, *P* = 0.031, Extended Fig. 3B,D), while media subclusters (K = 7, Extended Fig. 2C) were significantly associated with sex (AMI = 0.031, *P* = 0.040, Extended Fig. 3C,E). Subcluster assignments across subregions were neither identical nor mutually exclusive, suggesting that subregion heterogeneity emerges from distinct spatially organised biological processes.

**Figure 3.**
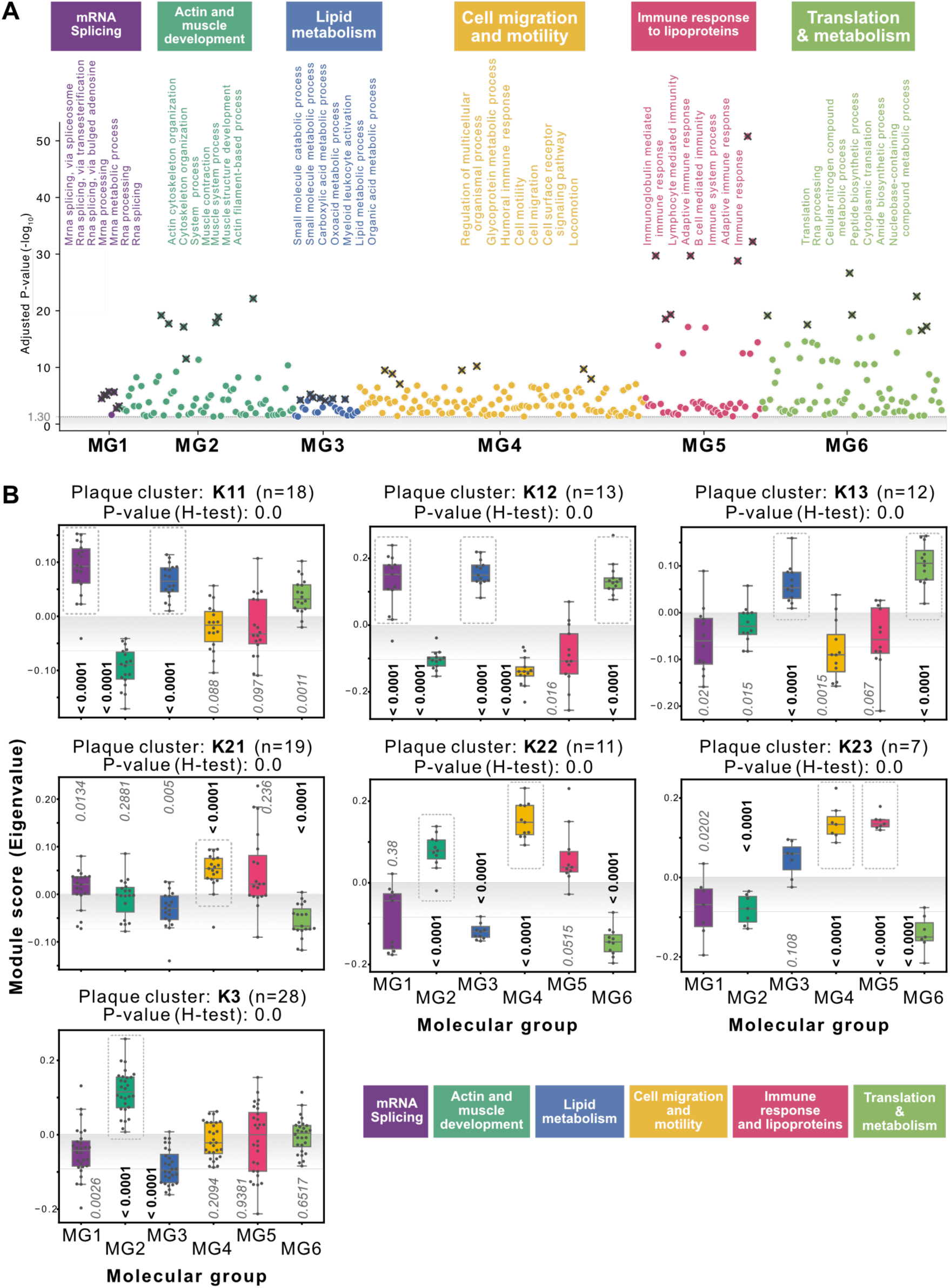
Necrotic core subtypes are distinctly associated with molecular group signatures. **A,** Manhattan plots showing Gene Ontology enrichment analysis of protein modules in necrotic core. **B**, Module score (eigenvalue) distributions across necrotic core clusters. Module scores represent the (correlation weighted) aggregate abundance of proteins in each molecular group. Dashed rectangles indicate statistically significant upregulation (p-value<0.05)

### Biological processes drive plaque subclusters

Next, we investigated the association between the plaque subclusters and biological processes. We grouped individual proteins into modules using weighted correlation network analysis (necrotic core: MG0–6, Fig. 2C, Extended Fig. 2D; fibrous cap and media: Extended Fig. 2E–F, Extended Fig. 3D–E). Each protein module was examined for overrepresented biological processes (Fig. 3A). For instance, in the necrotic core, MG1 was enriched for mRNA splicing and spliceosomes snRNP components, while MG2 encompassed actin cytoskeleton and muscle contraction pathways. The remaining protein modules showed enrichment in lipid metabolism (MG3), cell migration and motility (MG4), immune responses and lipoproteins (MG5), and translation/metabolism (MG6). We designated the protein modules using the top seven enriched biological processes. As expected, MG0 showed no distinct enrichment, reflecting the absence of consensus correlation among its constituent proteins. Protein modules in the fibrous cap and media shared similar functional themes but differed in size and number (Extended Fig. 3F-G).

Finally, to integrate these biological processes into a spatial context, we represented the abundance status of each process using module scores (Fig. 3B). Notably, all seven necrotic core subclusters had significantly distinct module profiles (*P* < 1 × 10^– 7, Kruskal–Wallis test), and each subcluster was associated with unique patterns of upregulated protein modules (*P* < 0.0001, one-sided bootstrap test). We observed distinct protein module patterns in the fibrous cap and media subregions, highlighting the spatial heterogeneity of biological processes in advanced (stable and vulnerable) plaques (Extended Fig. 4A–B)^13^.

**Figure 4.**
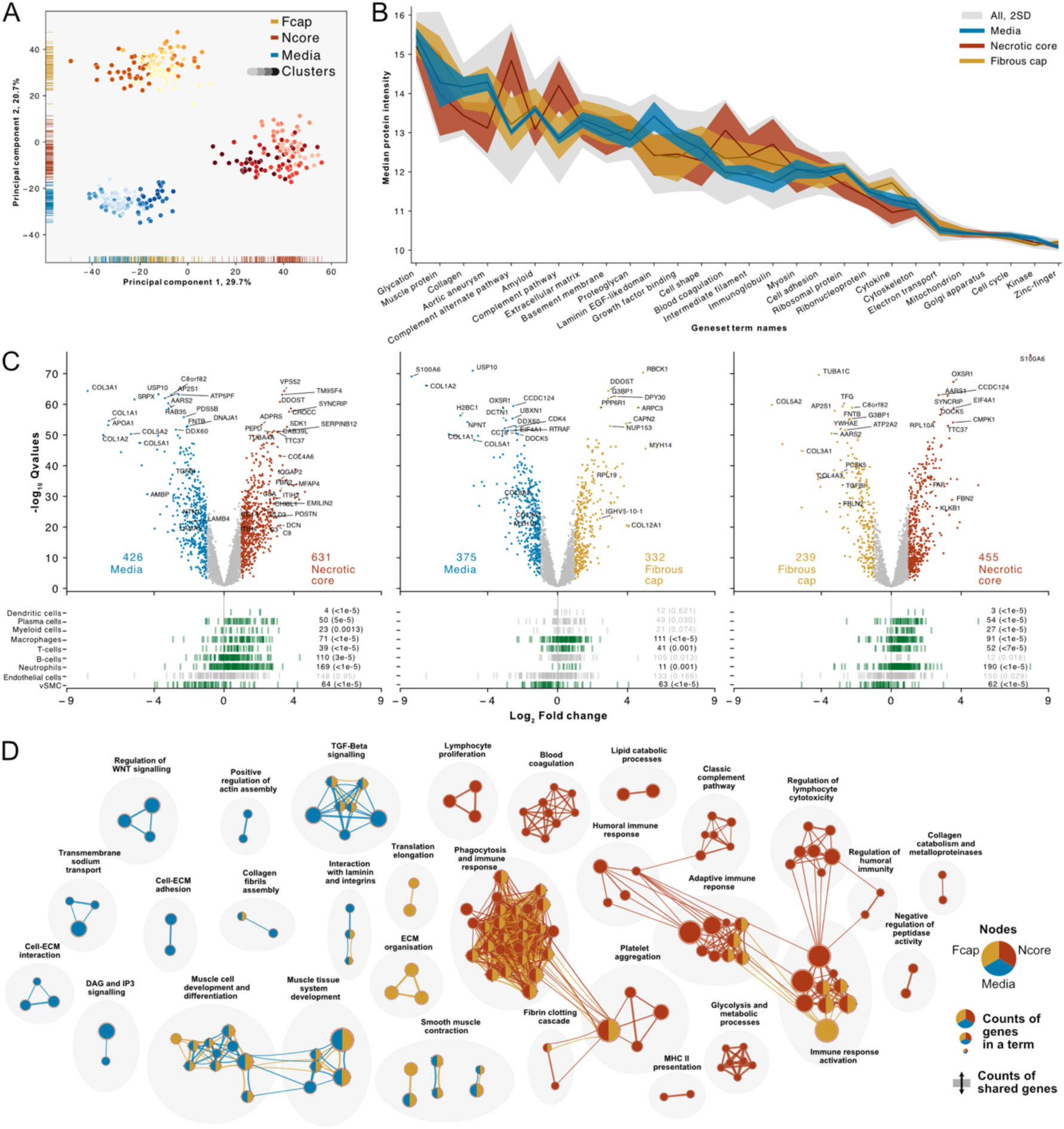
Molecular and cellular heterogeneity across atherosclerotic plaque subregions reveal specific biological processes. **a**, Principal component analysis of plaque proteomes (n=318 samples) demonstrates distinct quantitative profiles across histomorphological subregions (media, necrotic core, fibrous cap), with subregion clusters shown in varying shades. **b**, Protein abundance patterns across functional categories show region-specific enrichment. Lines indicate median abundance; shaded areas represent ±2SD. **c**, Differential protein analysis between paired regions reveals distinct molecular signatures (Q<0.05, |log2FC|>1). Cell-type deconvolution analysis (lower panels) demonstrates region-specific enrichment of cellular markers, with statistical significance determined by bootstrap testing. **d**, Network analysis of enriched biological processes highlights region-specific pathway activation. Connected nodes represent related pathways, revealing coordinated biological processes characteristic of each plaque region.

### Molecular and cellular characteristics of plaque subregions

Given the distinct histomorphological architecture of the three subregions, we investigated the associations between protein abundance patterns and spatial subregions. A variance-based analysis revealed the necrotic core was the most molecularly distinct subregion (Fig. 4A). The first variance dimension (PC1, 29.7% of total variance) separated the necrotic core (PC1 range: 20–60), while the second dimension (PC2, 20.7% of variance) distinguished the media (PC2 range: –20 to –40) and the fibrous cap (PC2 range: 20–40).

To pinpoint the biological mechanisms underlying these distinct profiles, we tested the associations between protein abundances and functional categories^22^ (Fig. 4B). In the media, highly abundant proteins were linked to muscle contraction, collagen, and proteoglycans (FDR < 0.01), highlighting its structural role and VSMC contractility. By contrast, immune and inflammatory pathways predominated in the necrotic core (FDR < 0.01). The fibrous cap showcased intermediate features, bridging the cell–ECM interactions of the media with the inflammatory characteristics of the necrotic core (FDR < 0.01). Notably, intracellular signalling and cytosolic proteins (e.g., mitochondrial, kinases, cell cycle factors) were uniformly low in abundance and variance.

Pairwise comparisons further highlighted subregion-specific protein abundance patterns (Fig. 4C, top). The necrotic core had the most distinctive profile, with 631 and 455 proteins upregulated relative to the media and fibrous cap, respectively (Q < 0.05). These included inflammatory and immune markers (e.g., CD1, ITIH2, C9) and ECM remodelling proteins (e.g., POSTN). By contrast, fewer proteins were uniquely upregulated in the media (375) and fibrous cap (332). Media-enriched proteins, such as COL3A1, COL1A2, and COL5A2 confirmed its role in arterial structure and VSMC function, whereas fibrous cap enriched proteins (e.g., COL12A1, RPL19) underscored its stabilising barrier role.

Inferred immune cell-type associations reinforced these functional distinctions (Fig. 4C, bottom). The necrotic core was enriched for macrophage, neutrophil, and other immune cell markers (P < 0.002), consistent with its prominent inflammatory profile. By contrast, the media was characterised by VSMC markers (P < 0.002), reflecting its contractile function in maintaining vascular integrity. The fibrous cap was enriched for immune cell and VSMC markers (P < 0.002), highlighting its intermediate role in structural stability and immune modulation.

Finally, pathway enrichment analyses confirmed the alignment between the molecular profiles and the histomorphological features of each subregion (Fig. 4D). The necrotic core was enriched for inflammation, immune activation, and catabolic processes, alongside pathways related to platelet aggregation and fibrin clotting—highlighting its thrombogenic potential. The media was predominantly enriched for smooth muscle cell contraction, muscle cell development, and cell–ECM interactions, reflecting its critical role in vascular structure and contractility. Meanwhile, the fibrous cap had a mixed profile, sharing features of the media and necrotic core, including ECM remodelling, VSMC differentiation, structure, and inflammation processes.

### Contrastive analysis of stable and vulnerable plaques

Having established the link between spatial subregions and distinct biological processes, we next examined how clinical covariates correlate with protein abundances in stable and vulnerable plaques. We observed a robust association between subregions and clinical covariates (Fig. 5A). In the necrotic core, 454 proteins were significantly associated with vulnerability status and 17 with rupture status (Q < 0.05). By comparison, 98 proteins in the fibrous cap were associated with vulnerability.

**Figure 5.**
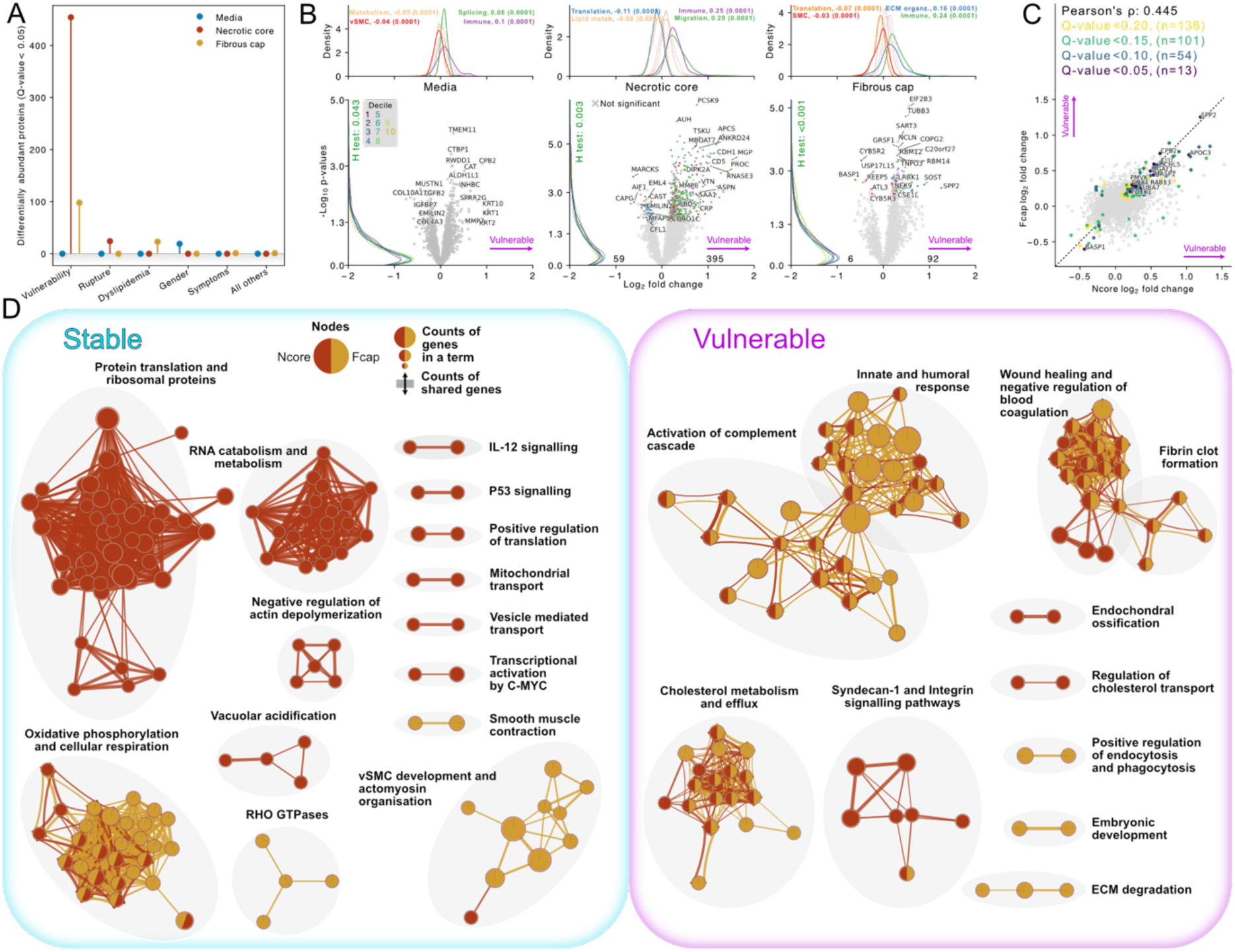
Subregion specific molecular signatures of plaque vulnerability. **a**, Counts of proteins significantly associated (q-value < 0.05) with clinical covariates, assessed by moderated t-test analysis. **b**, Protein abundance differences between stable and vulnerable plaques across three subregions: media (left), necrotic core (center), and fibrous cap (right). Lower panels show individual protein changes, with numbers indicating significantly differential proteins (q-value < 0.05, robustness score > 0.95) in stable and unstable plaques. Significantly differential proteins are coloured by protein module. Upper panels display density distributions of proteins in the protein modules with their median log2 fold changes, and bootstrap-derived p-values in parenthesis. **c**, Correlation of vulnerability-associated protein changes between necrotic core and fibrous cap (Pearson’s ρ=0.445), with proteins coloured by statistical confidence thresholds. **d**, Network analysis of enriched biological processes reveals distinct pathway activation patterns in stable (left) versus vulnerable (right) plaques.

Notably, the necrotic core had the most distinctive vulnerability-associated proteome, with 395 proteins upregulated (FDR < 0.05, Fig. 5B, bottom centre). Among these were MGP (up 1.5-fold) and AHSG (up 0.9-fold), supporting the concept of increased ossification and microcalcification in vulnerable plaques. Cholesterol transport and processing proteins were also increased, including APOC3 (1.3-fold), APOA5 (0.8-fold), and PCSK9 (0.7-fold), underscoring the role of lipid dysregulation in plaque vulnerability. Moreover, ECM remodelling proteins were elevated (e.g., GAS6, ACAN, and DPT), highlighting the active-matrix degradation and tissue remodelling processes. Immune response– and cell migration—related protein modules were similarly upregulated (P < 0.0001, Fig. 5B, top-centre).

In the fibrous cap, proteomic changes bridged structural features of the media with inflammatory and matrix-remodelling pathways characteristic of the necrotic core (Fig. 5B, bottom right). Compared to stable plaques, vulnerable lesions had greater abundances of ECM-destabilising proteins (SOST, TUBB3, REEP5) and immune modulators (LRRK1, NEK9). In parallel, ECM reorganisation and immune processes protein modules were significantly upregulated (P < 0.0001, Fig. 5B, top right), whereas protein translation and smooth muscle–associated modules were downregulated. By contrast, the media had comparatively modest abundance differences when comparing stable to vulnerable plaques (Fig. 5B, bottom left). Cytokeratin (KRT10, KRT1, KRT2) and cell differentiation markers had the largest shifts, reflecting VSMC dysfunction and phenotypic switching that accompany medial remodelling. However, these changes did not reach statistical significance (FDR > 0.05).

Correlation between necrotic core and fibrous cap vulnerability changes revealed 13 proteins with pleiotropic functions (Pearson’s ρ = 0.45, Q < 0.05, Fig. 5C). Notably, SPP2 (involved in calcification and matrix remodelling) was elevated by 1.2-fold in both subregions. Protein interaction analyses revealed a coordinated network involving calcification (SPP2), thrombosis regulation (CPB2), lipid metabolism (APOC3), cytoprotection (CLU), and complement-driven inflammation (MASP2) (Extended Fig. 8). We further investigated the spatial coordination by comparing protein–protein correlations across subregions. The necrotic core–fibrous cap interface had the most significant increase in correlation regarding vulnerability status (ρ_stable_ = 0.41 vs. ρ_vulnerable_ = 0.45, P = 2.91 × 10^–89, Extended Fig. 5A). Immune response–ECM remodelling, cell migration–immune activation, and mRNA splicing were the most significantly coordinated protein modules in vulnerable lesions (P = 5.1 × 10^–58, chi-square test; Extended Fig. 5B, C), underscoring once more the importance of spatially orchestrated inflammatory and structural changes in vulnerable and stable plaques.

Lastly, an integrative pathway enrichment analysis confirmed the distinct biological processes differentiating stable from vulnerable plaques (Fig. 5D). Stable plaques were enriched for cellular respiration (oxidative phosphorylation, FDR = 2 × 10^–4; aerobic respiration, FDR = 1.2 × 10^–3), active protein synthesis (cytoplasmic translation, FDR < 1 × 10^–5; cytoplasmic ribosomal protein, FDR < 1 × 10^–5), and smooth muscle cell functions (muscle cell development, FDR = 1.1 × 10^–3; smooth muscle contraction, FDR < 1 × 10^–5). These findings underscore the relevance of active metabolism, robust protein synthesis, and vigorous VSMC function in plaque stability. In contrast, vulnerable plaques were enriched for immune responses, cholesterol metabolism, wound healing, ossification, and ECM degradation pathways, as well as Syndecan-1 and integrin signalling. Collectively, these results highlight the pivotal roles of inflammation, lipid dysregulation, VSMC dysfunction, and ECM remodelling as key drivers of plaque vulnerability^16^.

### Pro-inflammatory VSMCs secrete PCSK9

Having investigated the spatial selectivity of vulnerability-related processes, we next investigated the mechanistic link between plaque vulnerability status and local PCSK9 abundance. We focused on VSMCs because they produce and secrete arterial PCSK9^23^, are involved in multiple vascular pathologies (aortic aneurysms, hypertension, Moyamoya disease), and exhibit disease-triggering phenotypic plasticity in response to local stressors^24–28^. We hypothesised that the oxidative and inflammatory microenvironment of the necrotic core not only promotes a pro-inflammatory VSMC phenotype, but also drives local PCSK9 upregulation, thereby contributing to the inflammatory, lipid-processing, and ECM-degrading processes characteristic of vulnerable plaques. To test this hypothesis, we recapitulated the oxidative and inflammatory conditions of the necrotic core in primary VSMC in vitro cultures via stimulation with POVPC (1-palmitoyl-2-[5-oxovaleroyl]-sn-glycero-3-phosphocholine), a pro-inflammatory oxidised phospholipid abundant in advanced atherosclerotic lesions (Fig. 6A)^29–31^. We assessed the functional role of PCSK9 in POVPC-activated VSMCs by transcript knockdown using Inclisiran (lacking its N-acetylgalactosamine moiety), a clinically approved siRNA that targets hepatocytic PCSK9 in patients with primary hypercholesterolemia or mixed dyslipidemia^32^.

**Figure 6.**
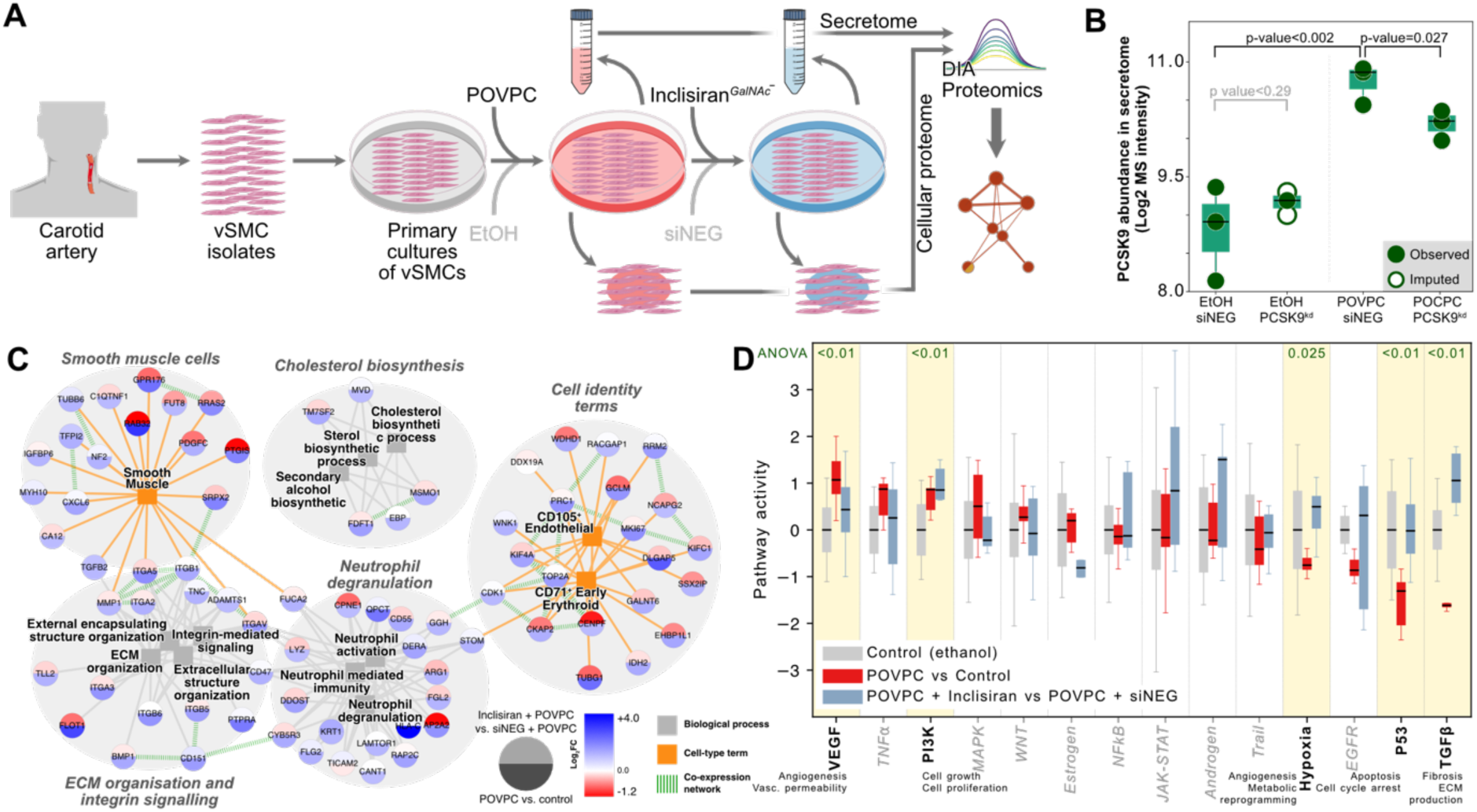
Functional validation of vulnerability mechanisms in vascular smooth muscle cells. **A**, Experimental workflow for characterising inflammatory and oxidative stress in VSMCs. **B**. PCSK9 protein abundance (log2 MS intensity) in observed (dark dots) and imputed (light dots) samples across conditions, with statistical comparisons shown. **C**, Knowledge-graph analysis of POVPC-induced gene expression changes; **D**, Pathway activation analysis comparing control (ethanol, grey), POVPC treatment (red) and PCSK9 knockdown (Inclisiran, blue) effects.

Our proteomic assays quantified an average of 7,563 cellular and 2,345 secreted proteins (Extended Fig. 6A). Notably, PCSK9 abundance increased 1.8-fold following POVPC treatment (P < 0.002, two-sided t-test) and decreased by 0.5-fold upon treatment with Inclisiran (P = 0.027, Fig. 6B). Among the experimental conditions, POVPC treatment drove the largest proteomic changes (748 proteins upregulated, FDR < 0.05, Extended Fig. 6C, D). Notably, neutrophil degranulation, ECM organisation, integrin signalling, and cholesterol biosynthesis pathways were all upregulated in POVPC-treated VSMCs (Fig. 6C). In contrast, PCSK9 knockdown in POVPC-treated VSMCs increased cell development processes and enhanced negative regulation of cytokine production (Extended Fig. 6E, F).

We further assessed the cellular response induced by POVPC and PCSK9 knockdown using pathway activity^33^ analysis (Fig. 6D). POVPC treatment promoted VSMC dysfunction by activating angiogenic (VEGF) signalling, while suppressing DNA damage responses (P53) and ECM production (TGFβ). In contrast, Inclisiran-mediated PCSK9 knockdown restored VEGF, hypoxia, and P53 signalling, and further enhanced cell growth (PI3K) and ECM production (TGFβ). Together, these findings confirm that oxidative and inflammatory stress drive a pro-inflammatory, atherogenic VSMC phenotype, marked by increased PCSK9 expression. Conversely, inhibiting PCSK9 drives a shift toward an atheroprotective, matrix-synthetic phenotype, marked by enhanced proliferation and ECM production. Altogether, direct arterial targeting of PCSK9 may offer novel therapeutic avenues for attenuating plaque vulnerability as well as other vascular pathologies, extending beyond the established lipid-lowering role of hepatocyte-derived PCSK9^23,34^.

### Prioritising proteins for predicting plaque vulnerability

Having shown that our proteomics approach can pinpoint spatially confined changes in stable and vulnerable plaque composition, we next sought to develop a protein biomarker panel to predict lesion stability status. Given the hierarchical nature of our data, we implemented a comprehensive strategy that integrates plaque subregion and subcluster annotations for prediction robustness. Our approach uses statistical methods to maximise biological effect sizes and metaheuristic optimisation with ensemble learning to refine the protein panel (Fig. 7A).

**Figure 7.**
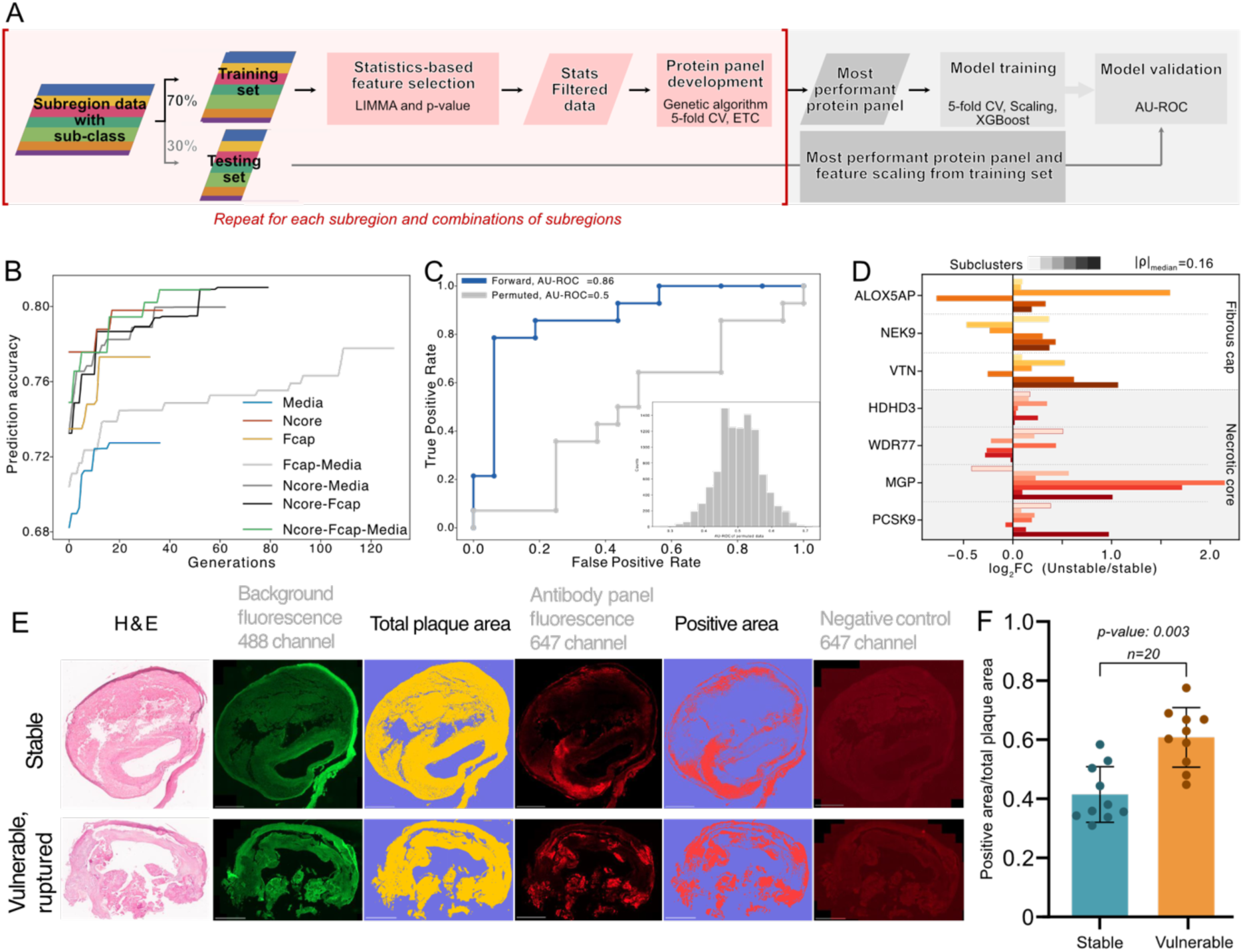
Development and validation of a protein signature for predicting plaque vulnerability. **a**, Machine learning workflow for identifying predictive protein markers from plaque proteomes. A genetic algorithm with cross-validation identified optimal protein combinations from training data (70%) and validated them in an independent test set (30%). **b**, Optimization of prediction accuracy across algorithm generations shows superior performance when combining multiple plaque regions. **c**, The optimised protein panel achieves robust prediction of plaque vulnerability (AUC=0.86), significantly outperforming random models (inset: permutation analysis, n=50,000). **d**, Expression patterns of the seven-protein signature across plaque regions and vulnerability states reveal consistent changes (median |ρ|=0.16). **e**, Validation of the protein signature by multiplexed immunofluorescence in human plaques. Representative images show increased signature proteins in vulnerable versus stable plaques. Individual scans are available (Extended Fig.9 A stable, B vulnerable). **f**, Quantification confirms significantly higher protein panel expression in vulnerable plaques (p=0.003, n=20, two-tailed t-test).

**Figure 8.**
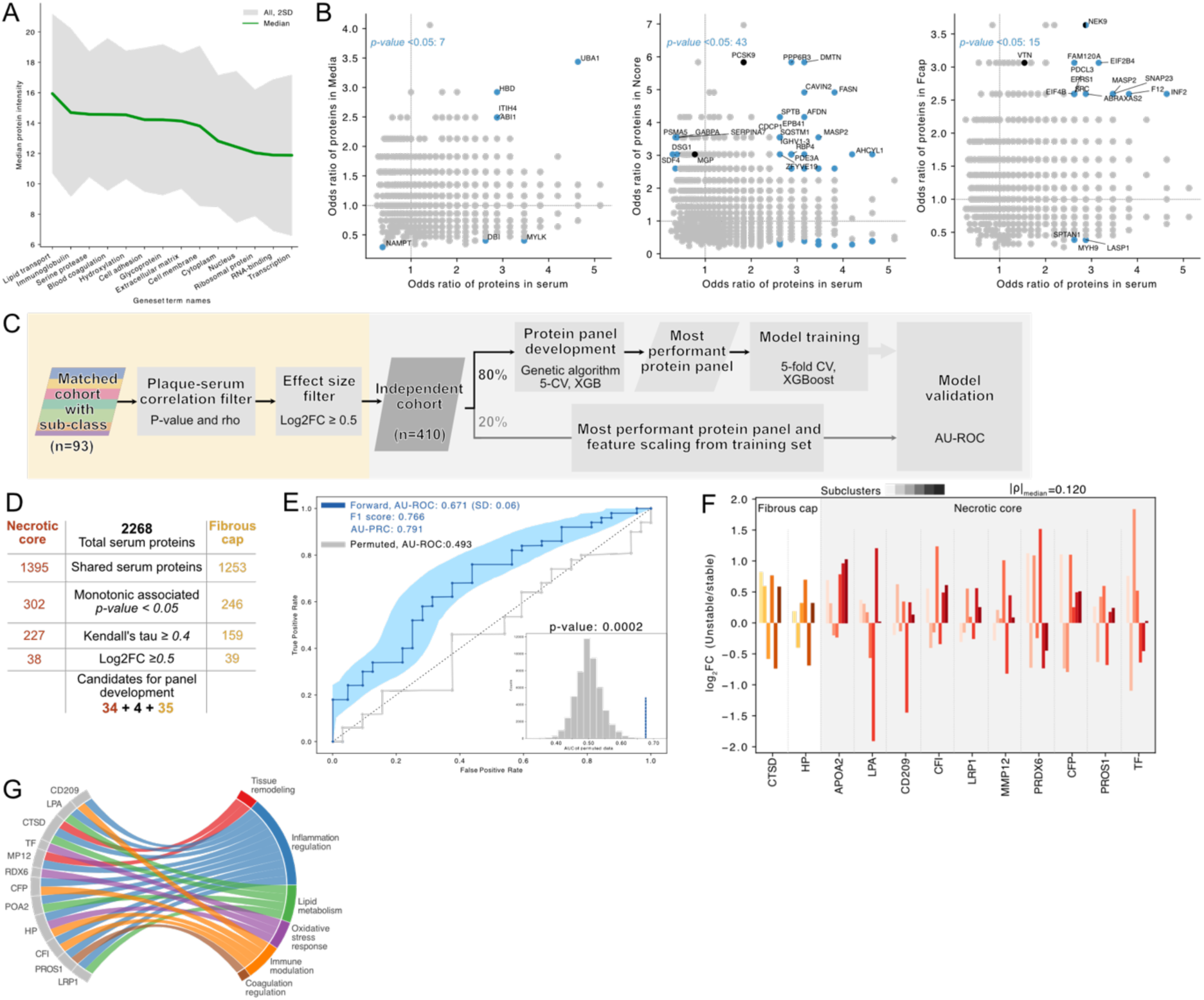
Identification of serum protein signatures for non-invasive detection of plaque vulnerability. **a**, Serum protein abundance distribution across functional categories, showing median protein intensities (green line) with biological variation represented by ±2 standard deviations (grey band). **b**, Correlation analysis of protein enrichment (odds ratios) between serum and plaque subregions: media (left), necrotic core (middle), and fibrous cap (right). Odds ratios >1 indicate protein enrichment in vulnerable plaques. Blue dots represent proteins with significant associations in both serum and respective plaque regions (Fisher’s exact test, p < 0.05). Black dots indicate proteins from the previously optimised plaque tissue panel. **c**, Serum biomarker panel development workflow. Initial protein filtering was performed using the matched serum-plaque cohort, applying monotonic correlation and effect size thresholds for each plaque cluster (yellow, n=93). Filtered proteins were used for optimising protein panel in an independent cohort of serum samples (grey, n=410). **d**, Summary of the stepwise filtering process of serum proteins. **e**, Receiver operating characteristic (ROC) performance of serum protein panel (blue line, AUC = 0.67) compared to label permuted control (grey line, AUC = 0.50). The inset histogram shows the distribution of AUC values from 50,000 label permutation trials. **f**, Differential abundance patterns of the 12 proteins in the optimised serum panel in the matched (serum-plaque) cohort. Shading indicates corresponding subregion cluster and the proteins have a median absolute Pearson’s rho of 0.12. g, Functional description of the 12 serum proteins in the panel (left) with their associated biological processes (right).

This integrated approach revealed a hierarchical pattern consistent with our earlier findings. A marker panel derived from necrotic core proteins offered the highest individual predictive power (AU-ROC = 0.79), followed by panels from the fibrous cap (AU-ROC = 0.77) and the media (AU-ROC = 0.73) (Fig. 7B). Notably, combining proteins from multiple subregions improved performance further (AU-ROC = 0.82). An optimised seven-protein panel from the necrotic core and fibrous cap had the highest predictive plaque vulnerability performance (AU-ROC = 0.86, *P* < 0.0001, permutation test; Fig. 7C).

This optimised panel encompasses proteins with diverse functional roles in carotid artery disease. In the fibrous cap, ALOX5AP drives leukotriene-mediated inflammation and macrophage activity; NEK9 regulates microtubule dynamics, endothelial integrity, and apoptosis; and VTN governs ECM stability and thrombosis. From the necrotic core, HDHD3 participates in nucleotide metabolism and oxidative stress responses; WDR77 modulates gene expression and smooth muscle cell phenotypes; MGP promotes vascular calcification; and PCSK9 contributes to lipid metabolism and oxidative stress.

The low median correlation among these proteins (Absolute Pearson’s ρ = 0.16) underscores their complementary biological processes, thus enhancing the panel’s robustness (Fig. 7D). To validate the predictive performance, we performed quantitative immunofluorescence in an independent cohort of stable and vulnerable plaques (n=20, Fig. 7E). This confirmed the panel’s utility (*P* = 0.003), highlighting the potential for integrating multi-subregion proteomics into plaque risk stratification (Fig. 7F).

### Plaque vulnerability markers in serum proteome

Having identified vulnerability-associated proteomic signatures in plaque tissues, we next investigated whether signatures manifest in serum. We profiled serum proteomes of 503 CEA patients with stable and vulnerable carotid plaques (Table 1) and quantified 2,268 proteins using the perchloric acid–based enrichment method^35^ for serum proteomics. In contrast to plaque tissue proteomes, the most abundant serum proteins were lipid transport proteins, immunoglobulins, and serine proteases, whereas lower-abundance proteins included cytoplasmic, nuclear, and transcriptional regulators (Fig. 8A). This abundance gradient—from classical serum components to “leakage” proteins of tissue origin—illustrates the selective compartmentalisation of the serum proteome^36^.

The necrotic core had the strongest association with serum proteins (43 vs. 15 for the fibrous cap and 7 for the media, Fig. 8B). We adapted our plaque-based biomarker approach for serum analysis to prioritise proteins that predict vulnerability status (Fig. 8C). In the initial protein selection, we used a subset of cases with matched plaque and serum samples (n=93), while reserving the remaining 410 samples for optimisation and validation of vulnerability signature proteins. We filtered 2,268 serum proteins based on monotonic associations with plaque proteins (Extended Fig. 7B-C) and effect sizes for vulnerability (Extended Fig. 7B), narrowing the set to 38 necrotic core–associated and 39 fibrous cap–associated candidates (Fig. 8D, Extended Fig. 7D). A genetic algorithm–based optimisation then produced a 12-protein panel with an AU-ROC of 0.67 (p = 0.0002, permutation test; Fig. 8E).

This optimised panel has distinct subregion-specific abundance patterns and subcluster associations (median Pearson ρ = 0.120, Fig. 8F). Functional annotation showed enrichment of processes central to plaque vulnerability, including inflammation regulation (CD209, HP, CFI), tissue remodelling (CTSD, MMP12), lipid metabolism (APOA2, LPA), oxidative stress response (PRDX6, TF), and coagulation regulation (PROS1, Fig. 8G). These findings highlight the inflammatory, lipid-processing, and tissue-remodelling pathophysiology underlying vulnerable plaques and suggest that a serum-based signature may provide a non-invasive vulnerability assessment.

## Discussion

Our study of CEA patients with advanced carotid plaques demonstrates the relationship between subregion-confined biological processes and plaque vulnerability. It supports the concept that plaque composition, rather than stenosis severity, governs plaque vulnerability^37,38^. Using high-resolution mass spectrometry, we profiled the spatial proteomes of stable and vulnerable carotid plaques from symptomatic and asymptomatic patients. Our findings showed that the necrotic core has the most distinct vulnerability-related proteomic profile as it had the largest number of differentially abundant proteins. Importantly, we identified candidate biomarkers— in plaque tissue and serum—that distinguish stable from vulnerable plaques. A seven-protein panel derived from the necrotic core and fibrous cap had the highest predictive accuracy. Furthermore, functional experiments confirmed that oxidative and inflammatory stimuli upregulate PCSK9 expression in VSMCs, thereby highlighting a mechanistic link between plaque vulnerability and VSMC phenotypic modulation and plasticity^39^.

A principal finding of this work is the stark difference in the necrotic core proteomes of stable and vulnerable plaques. The upregulation of inflammatory mediators, lipid-transport proteins, and ECM-remodelling factors in the necrotic core aligns with previous evidence implicating local inflammation and lipid dysregulation in plaque rupture^40,41^. We expand on these findings by identifying specific functional processes —particularly ossification, immune cell infiltration, and lipid processing—that enhance plaque vulnerability^42,43^. The observation that multiple molecules (e.g., MGP, PCSK9) are consistently elevated in vulnerable lesions emphasises the role of chronic inflammation and abnormal lipid trafficking in vulnerable plaques.

Although the fibrous cap is regarded as the static structural barrier preventing rupture, we uncovered unexpected proteomic changes indicating significant upregulation of inflammatory, cholesterol metabolism, and ECM-remodelling processes. Several proteins known for modulating ECM integrity (e.g., SOST, TUBB3) were upregulated in vulnerable fibrous caps, suggesting that the fibrous cap is not merely a passive barrier but instead undergoes active, localised remodelling that can either reinforce or erode plaque stability. These findings bolster emerging views that fibrous cap thinning is a dynamic, multifactorial process driven by inflammatory cells, VSMC plasticity, and alterations in ECM components^44^. Finally, although the media appeared less altered in vulnerable plaques, it showed markers indicative of VSMC dysfunction^45–47^.

Our study directly connects the oxidative and inflammatory microenvironment of the vulnerable necrotic cores and increased arterial PCSK9 abundance. The treatment with pro-inflammatory phospholipid POVPC promotes a pro-atherogenic senescent phenotype in primary cultures of VSMCs, as evidenced by the upregulation of VEGF signalling and neutrophil degranulation processes. Interestingly, siRNA-mediated knockdown of PCSK9 (using clinically approved Inclisiran) in the POVPC-activated VSMCs enhances atheroprotective processes (such as TGFβ signalling and downregulation of cytokine production). Furthermore, previous studies have shown that PCSK9 impairs cytokinesis and promotes polyploidisation—hallmarks of senescence^48^. The findings indicate that focusing on arterial PCSK9 may provide a new approach to reducing vulnerable plaques, additive to its established function of lipid metabolism in hepatocytes^49^.

Furthermore, we directly evaluated the translational aspects of our mechanistic findings. We developed predictive protein signature that effectively identified vulnerable plaques in tissue. The seven-protein panel derived from the necrotic core and fibrous cap had robust predictive accuracy, suggesting potential clinical utility for risk stratification in patients undergoing CEA^50^. Additionally, the modest performance of the serum protein-based signature reflects the inherent complexity of the serum proteome^50,51^. These outcomes emphasise that spatial, multi-subregion plaque proteomics can reveal distinct protein signatures and pathways for detecting plaque vulnerability.

Note that our study has limited applicability for early-stage atherosclerosis, as it could not capture the critical phase of neo-intima (early-stage necrotic core) formation from the media^52^. The cross-sectional design of this study provides only a single “snapshot” of protein dynamics, underscoring the need for longitudinal studies to correlate proteomic shifts with clinical outcomes such as plaque rupture. We deliberately chose carotid plaque vulnerability status as the surrogate endpoint rather than the combined endpoint of patient symptomatic status. Categorising plaques using a combined endpoint is inherently more complex and will introduce additional variability from the broad spectrum of initiators of clinical carotid artery disease symptoms (such as a transient ischaemic attack, amaurosis fugax, and stroke)^4,53,54^. Although a larger prospective cohort could stratify and resolve these complexities, our reliance on supervised histomorphology-based segmentation by pathologists restricts sample size. Consequently, an automated, AI-based plaque segmentation—validated against expert annotations—is necessary for larger-scale confirmation studies^55,56^. Finally, our in vitro findings suggest a link between plaque vulnerability, VSMC senescence, and PCSK9 abundance. However, follow-up studies with in-depth functional proteomics of PCSK9 (e.g., phosphoproteomics, protein-protein interaction analyses) are needed to confirm the underlying mutation, phosphorylation, and signal transduction mechanisms in greater detail^27,57–59^.

Despite the high prevalence of moderate stenosis, only a small proportion of patients experience transient ischaemic attacks or strokes. Performing CEA on all patients with moderate stenosis is impractical and unjustified, highlighting the need for biomarkers for risk stratification and a deeper understanding that links molecular features to plaque clinical outcomes. Through spatial proteomic analysis, our study addresses this need by demonstrating that lipid processing, ECM remodelling, inflammation, and ossification within the necrotic core and fibrous cap are critical drivers of carotid plaque vulnerability. The discovery that pro-inflammatory VSMCs actively secrete PCSK9 highlights a promising therapeutic target that can mitigate advanced-stage vulnerable plaques. As personalised cardiovascular medicine advances, integrating these proteomic signatures into clinical workflows can enhance risk stratification and guide more timely diagnostics and interventions. By offering further insight into the molecular foundations of plaque vulnerability, this study establishes a foundation for more nuanced, targeted research to improve patient stratification and reduce cerebrovascular events.

## Data Availability

All data produced in the present study are available upon reasonable request to the authors.

## Acknowledgement

We are grateful to all patients for their consent, which enabled this study. We also thank all members of the Department of Proteomics and Signal Transduction (Max Planck Institute of Biochemistry, Munich) and the Experimental Vascular Medicine Department (TUM, Munich). In particular, we thank S. Williems and C. Ammar for their fruitful discussions on optimising protein quantification and J. Wettich for his technical insights into implementing the genetic algorithm. Furthermore, we acknowledge the funding agencies that supported this study: the Bavarian Ministry of Health and Care through the research project DigiMed Bayern (to L.M., M.M., H.S., and M.vS.), along with the DFG-sponsored CRC1123 on ‘Atherosclerosis – Mechanisms and Novel Networks of Therapeutic Targets’ (to L.M., H.S.).

## Author contributions

A.S., N.S., E.K., and J.P. conducted the experiments and prepared the samples. N.S., L.M., and D.B. designed the cohort. E.K., S.S., T.N., and M.L.E. optimised and executed the laser microdissection. E.K., J.P., and H.W. carried out the in vitro experiments. E.K., S.S., V.A., P.S., and A.S. performed the mass spectrometry measurements. E.K. and V.A. conducted the serum proteomic experiments. N.S. and E.K. carried out the immunofluorescence experiments. A.S., T.N., E.U., and E.H.R. performed the statistical and bioinformatic analyses. M.v.S. and H.S. guided the conceptualisation and provided critical feedback on project milestones. A.S., N.S., L.M., and M.M. wrote the first draft of the manuscript. L.M. and M.M. secured the funding, supervised and directed the project, interpreted clinical and MS-based proteomics data, and authored the manuscript. All authors contributed feedback on the research, data interpretation, and article.

---

**Extended Fig. 1.**
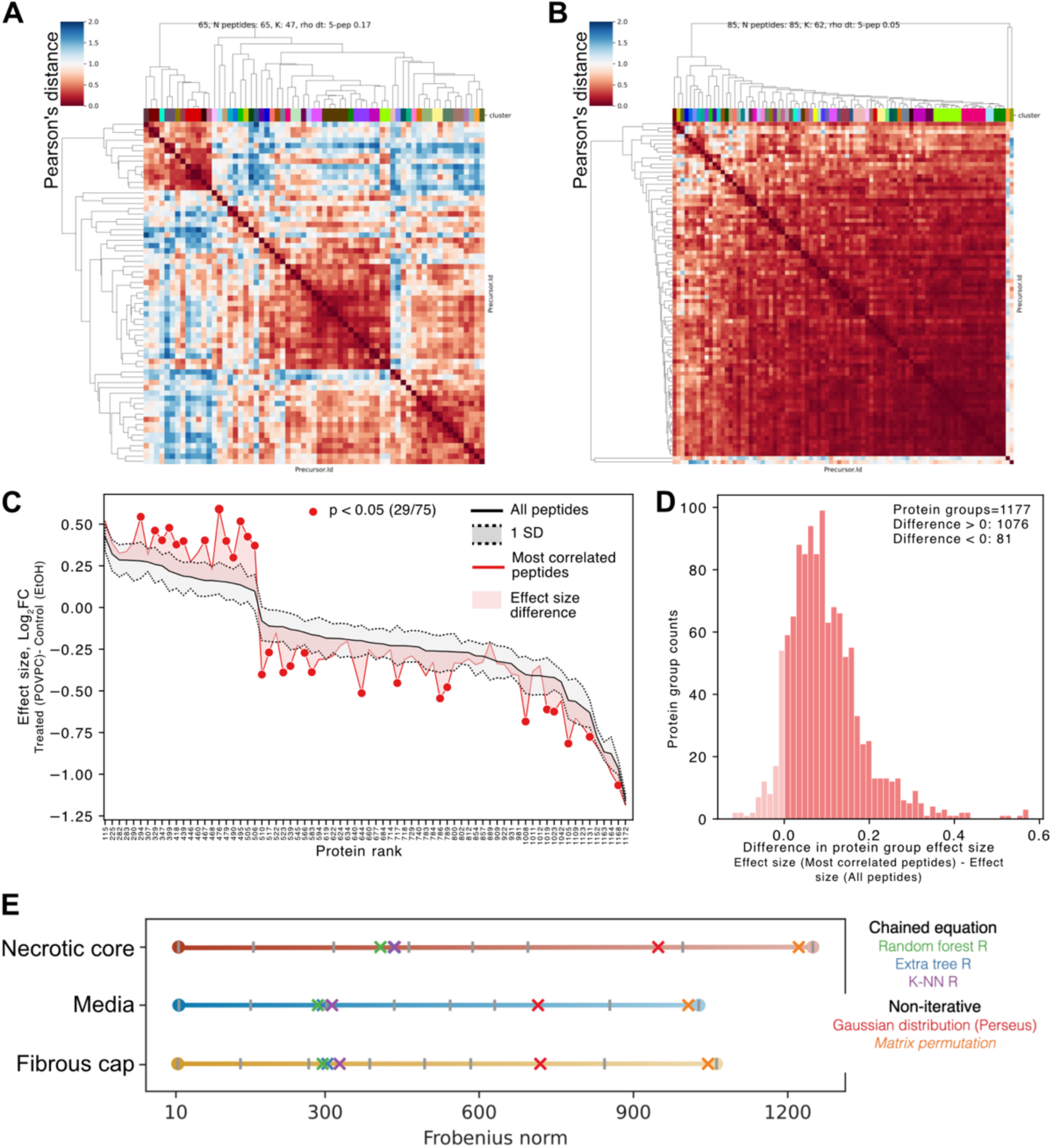
Overview of the refinement of the quantitative proteomics data. **A-B**, Internal consistency analysis shown by peptide correlation heatmaps for representative proteins: B, EEF2 (low consistency) and C, PPL (high consistency). Peptide clusters (K) were defined using weighted Pearson distance threshold (rho dt). **C**, Comparison of protein effect sizes calculated using all peptides (black) versus most correlated peptides (red) for selected significant proteins. Error bars represent standard deviation from bootstrap sampling (n=correlated peptide count, 1001 iterations). Twenty-nine proteins showed significantly different effect sizes. **D**, Distribution of effect size differences between analyses using most correlated versus all peptides, showing enhanced effect sizes in 1076 out of 1177 significant proteins. **E**, Evaluation of imputation methods using matrix (Frobenius) norm to quantify imputation artifacts. Abbreviations: CAD, coronary artery disease; BMI, body mass index; PAD, peripheral arterial disease; MI, myocardial infarction; AF, atrial fibrillation; COPD, chronic obstructive pulmonary disease; PFI, progression-free interval.

**Extended Fig. 2.**
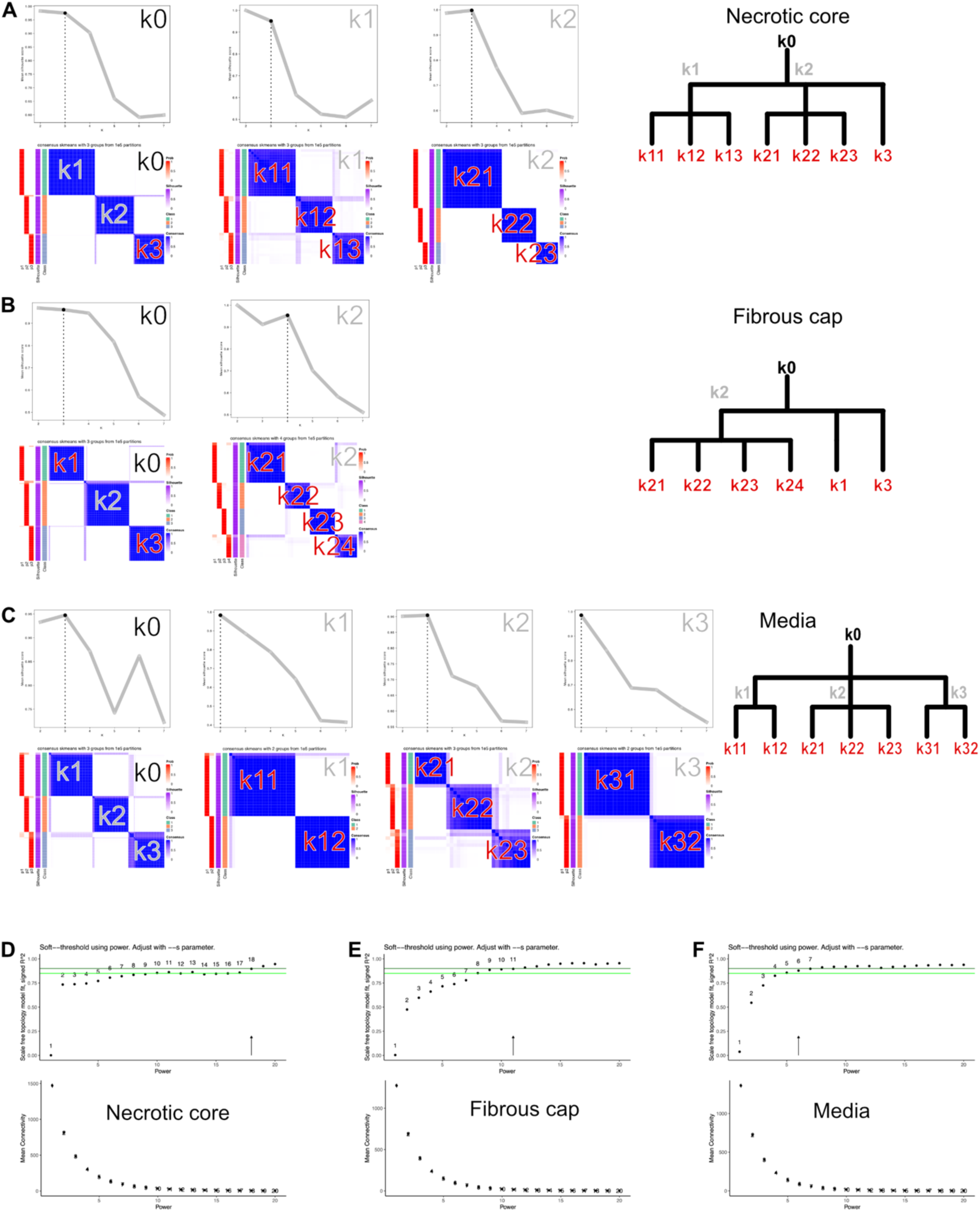
Identification of plaque tissue subclusters and protein modules. **A-C**, Hierarchical clustering analysis of distinct plaque regions: A, necrotic core; B, fibrous cap; and C, media. For each region, the dendrogram shows clustering hierarchy with root nodes (k0, black), internal nodes (grey), and final subclusters (red). Top panels: optimization of cluster number using silhouette scores. Bottom panels: consensus clustering matrices after 100,000 iterations. **D-E**, Weighted gene correlation network analysis (WGCNA) of proteins across the three plaque regions. Selection of optimal power parameter (β) to achieve scale-free topology (R² > 0.9, top). Module interconnectivity shows distinct protein modules at selected power (bottom).

**Extended Fig. 3.**
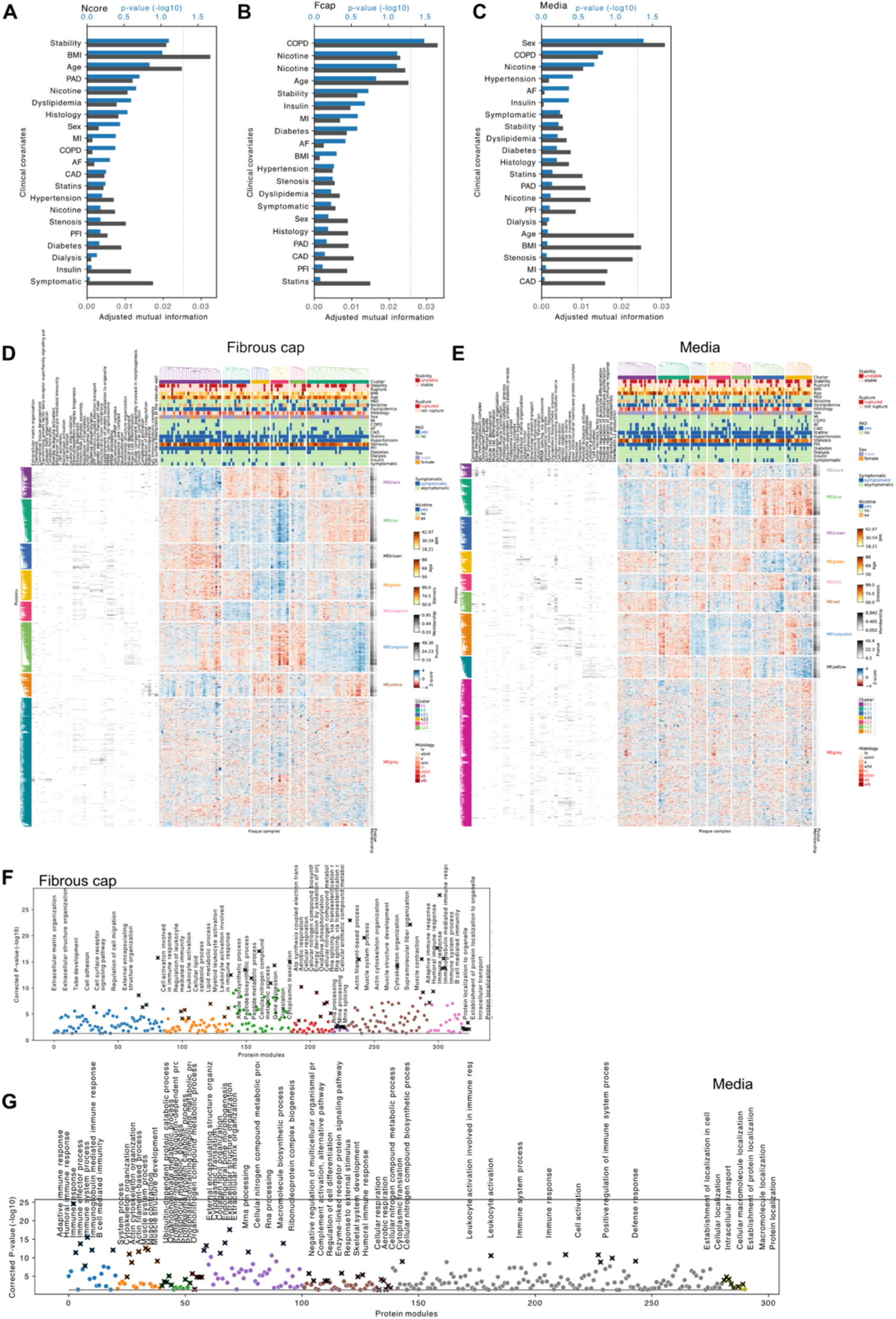
Clinical correlates and functional enrichment of plaque region subclusters. **A-C**, Association between clinical variables and identified subclusters in necrotic core (A), fibrous cap (B), and media (C). Significant associations were found only for COPD with fibrous cap clusters and sex with media clusters (p < 0.05). **D-E**, Consensus clustering analysis using the top 66% most variable proteins in fibrous cap (D) and media (E). **F-G**, Manhattan plots showing Gene Ontology enrichment analysis of protein modules in fibrous cap (F), and media (G). The seven most significantly enriched terms were used to annotate the protein module.

**Extended Fig. 4.**
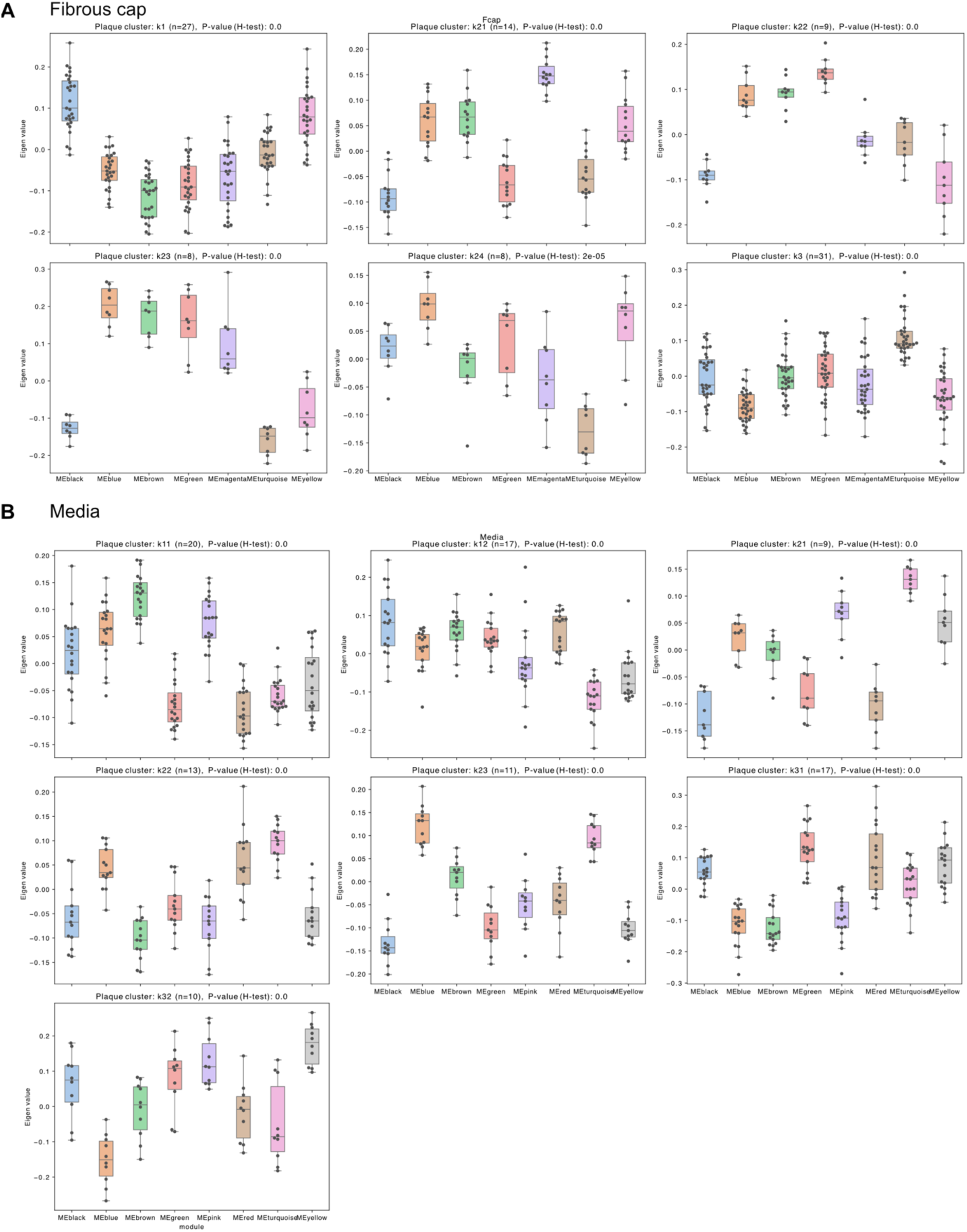
Distribution of molecular group signatures across plaque subclusters. **A**, Box plots showing molecular group scores (eigenvalues) for seven functional modules (colored bars) across six distinct plaque subclusters (k1-k24, k3) in the fibrous cap. Each subcluster panel shows sample size (n) and significance of module variation (H-test P-value). Individual data points are overlaid on boxes showing median, quartiles, and range. **B**, Similar analysis of molecular group scores across media subclusters.

**Extended Fig. 5.**
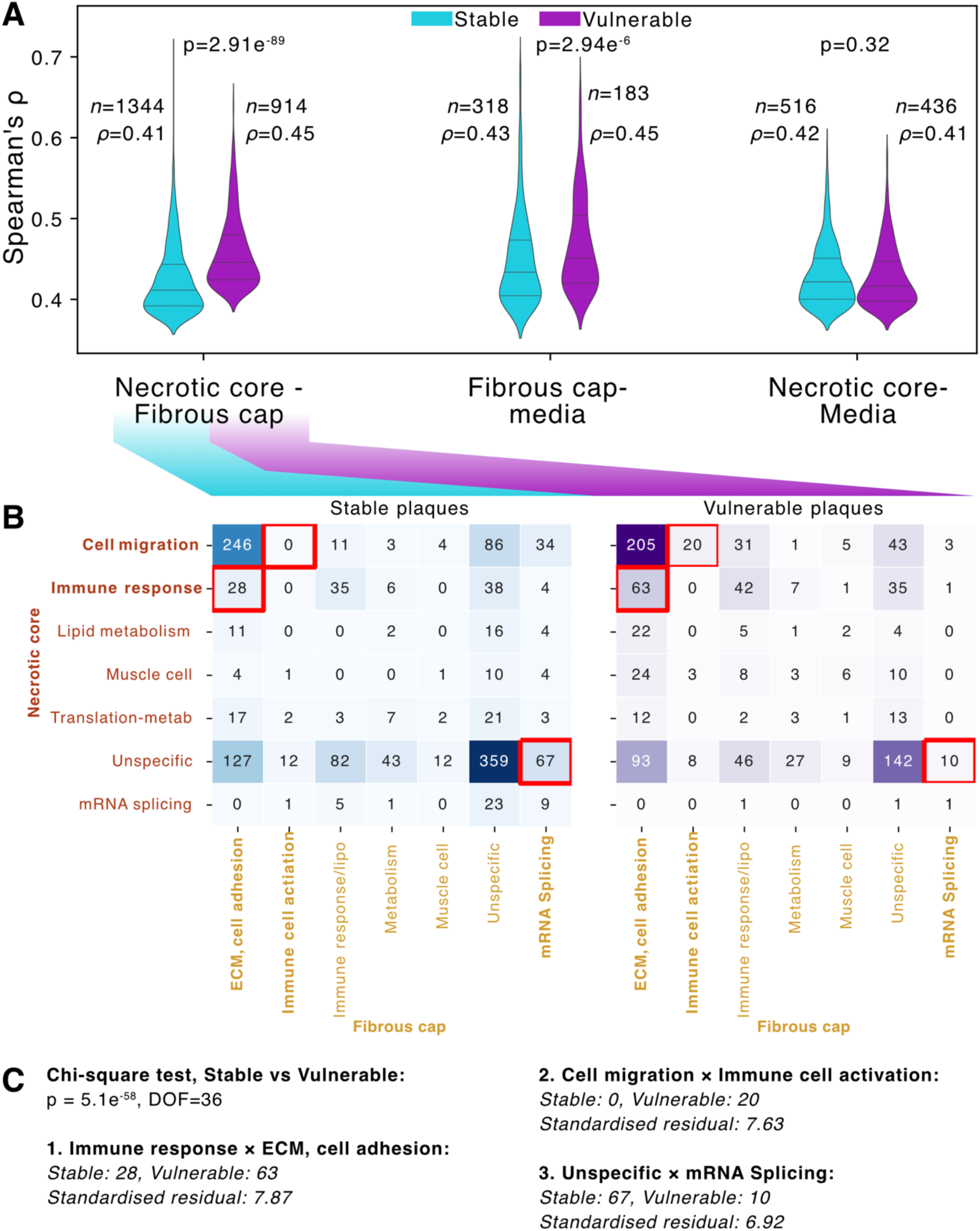
Protein correlation patterns between plaque regions in stable and vulnerable plaques. **A**, Violin plots showing distribution of Spearman’s correlation coefficients (ρ) between protein pairs across different plaque region comparisons. Sample sizes (n) and median correlation values are shown for each comparison. P-values indicate differences between stable and vulnerable plaques. **B**, Heat map showing the counts of correlated protein pairs between necrotic core (rows) and fibrous cap (columns) molecular groups in stable (left) and vulnerable (right) plaques. Numbers indicate protein pair counts. Red boxes highlight key differences between plaque types. **C**, Statistical comparison of protein pair frequencies between stable and vulnerable plaques. Overall difference was assessed by the Chi-square test, with three specific interactions showing the strongest standardised residuals.

**Extended Fig. 6.**
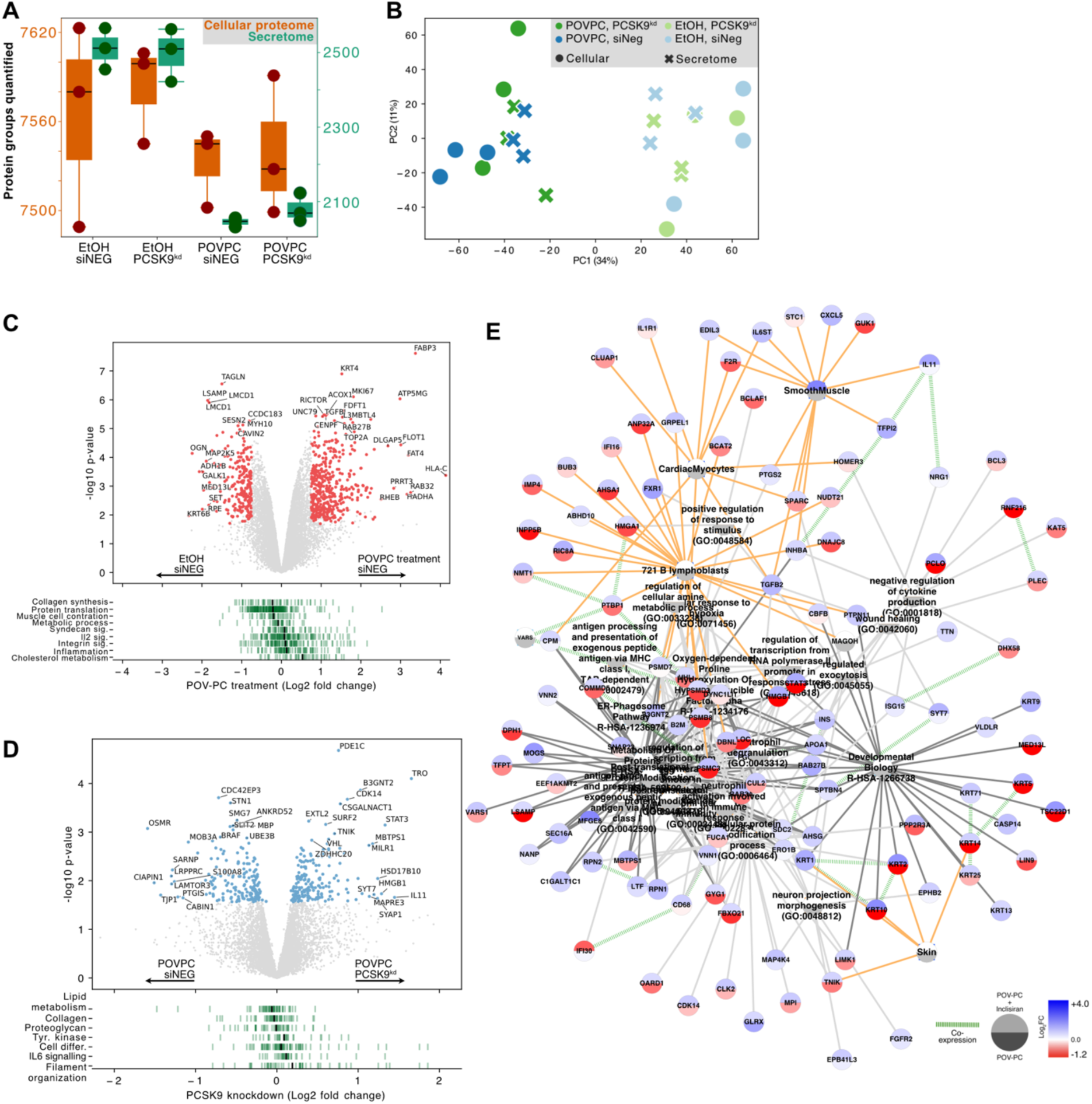
Proteomic analysis of POVPC treatment and PCSK9 knockdown in vascular smooth muscle cells. **A**, Number of proteins quantified in cellular proteome (orange) and secretome (green) across experimental conditions. **B**, Principal component analysis of cellular proteome and secretome samples showing separation of treatment conditions. **C-D**, Volcano plots showing differentially expressed proteins (red dots, p < 0.05) in response to D, POVPC treatment and E, PCSK9 knockdown compared to controls (siNEG). **E**, Integrated knowledge graph analysis of proteins significantly upregulated upon PCSK9 knockdown with Inclisiran (red: upregulated, blue: downregulated).

**Extended Fig. 7.**
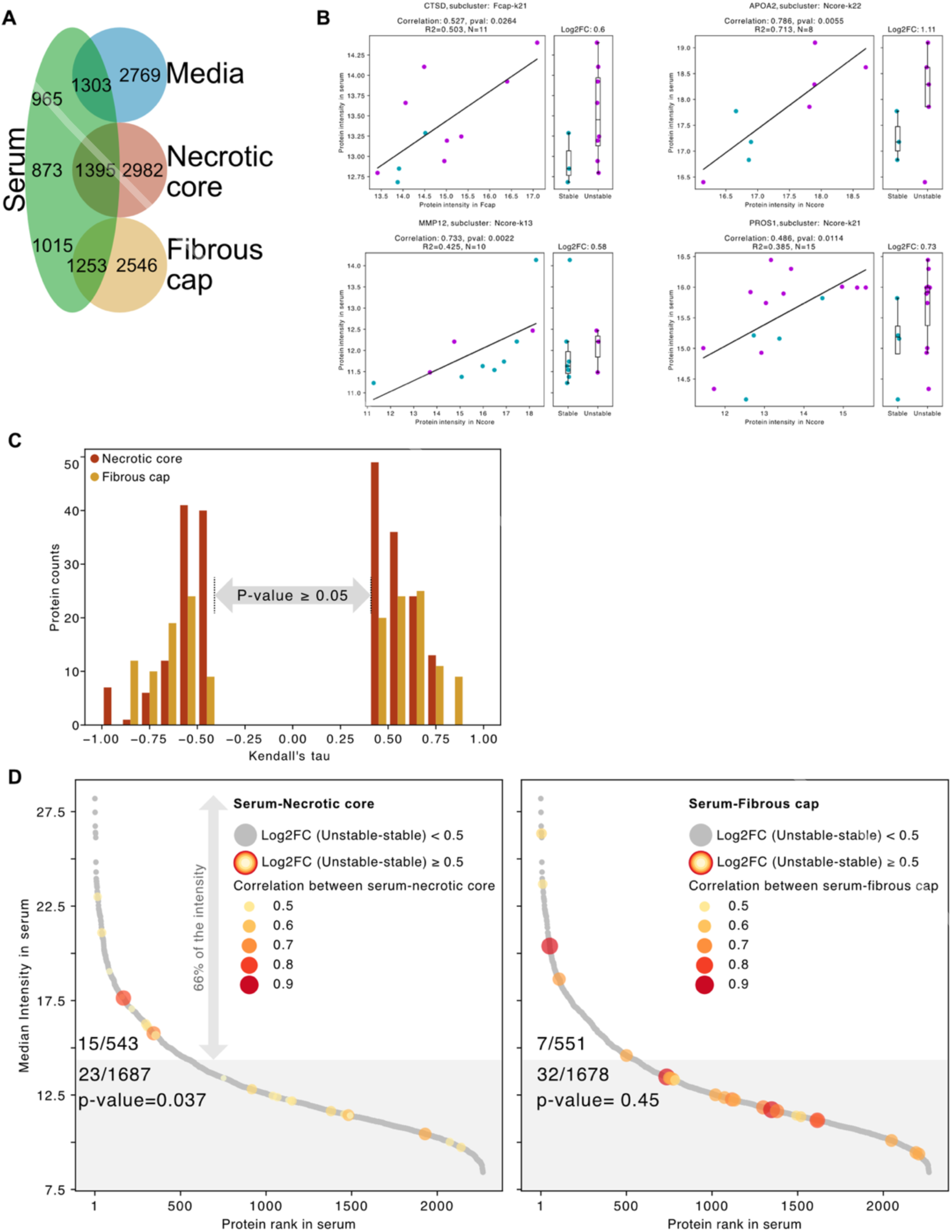
Selection of protein candidates for plaque vulnerability signatures in serum. **A**, Venn diagram showing the overlap of proteins identified across serum and three plaque regions. **B**, Correlation analysis of representative proteins between plaque regions and serum, showing protein intensities in stable (blue) and unstable (orange) plaques with corresponding box plots. Protein abundance correlation in matching tissue and serum are shown with Kendall’s tau correlation coefficients (R2). **C**, Distribution of Kendall’s tau correlation coefficients between serum and plaque protein levels in necrotic core (red) and fibrous cap (yellow), with regions with non-significant correlations (p ≥ 0.05) indicated. **D**, Ranked protein abundance in serum (grey dots) showing correlations with necrotic core (left) and fibrous cap (right). Colored dots indicate proteins with significant abundance changes between stable and unstable plaques (log2FC ≥ 0.5), with dot color intensity representing correlation strength. Numbers show proteins meeting both correlation and fold-change criteria.

**Extended Fig. 8.**
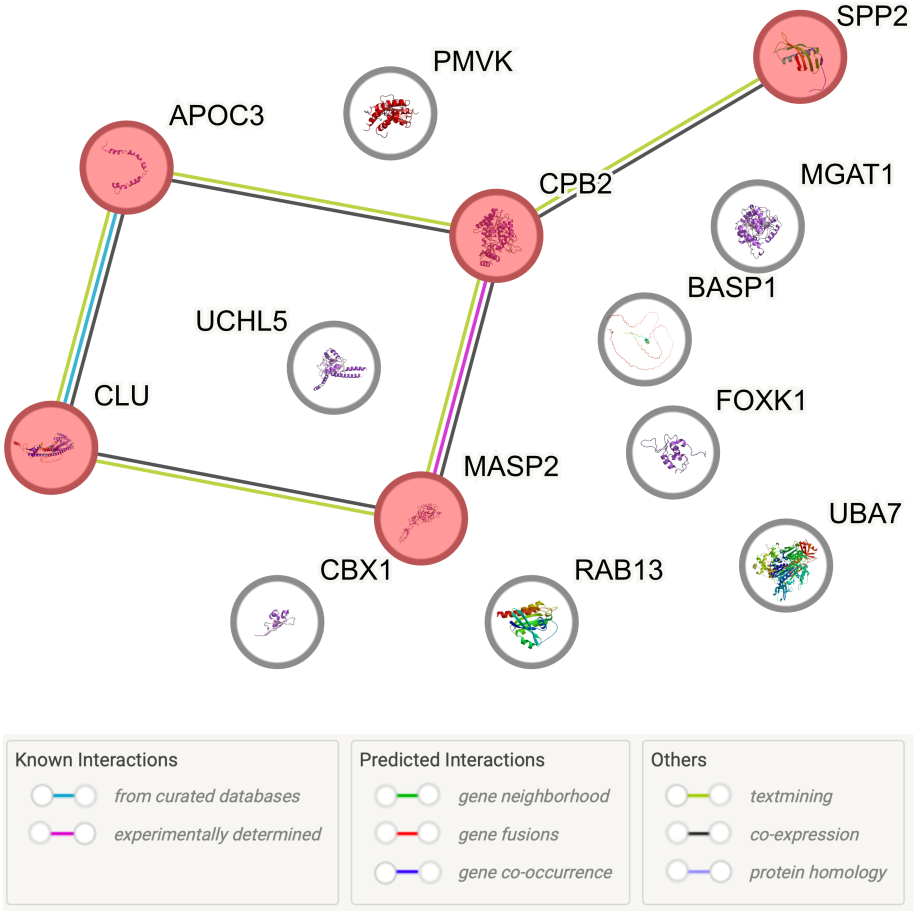
StringDB interaction analysis of correlated proteins.

**Extended Fig. 9.**
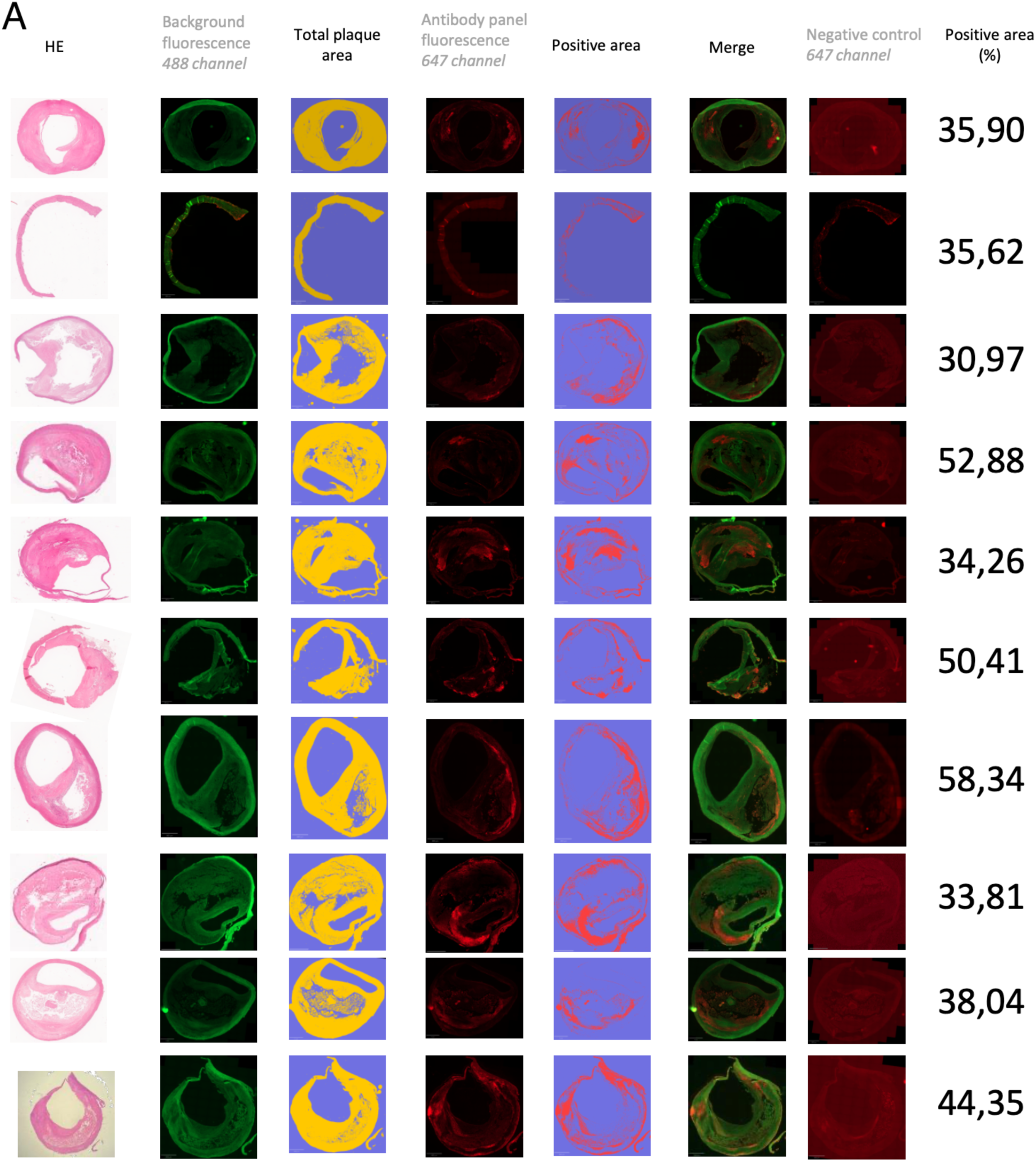
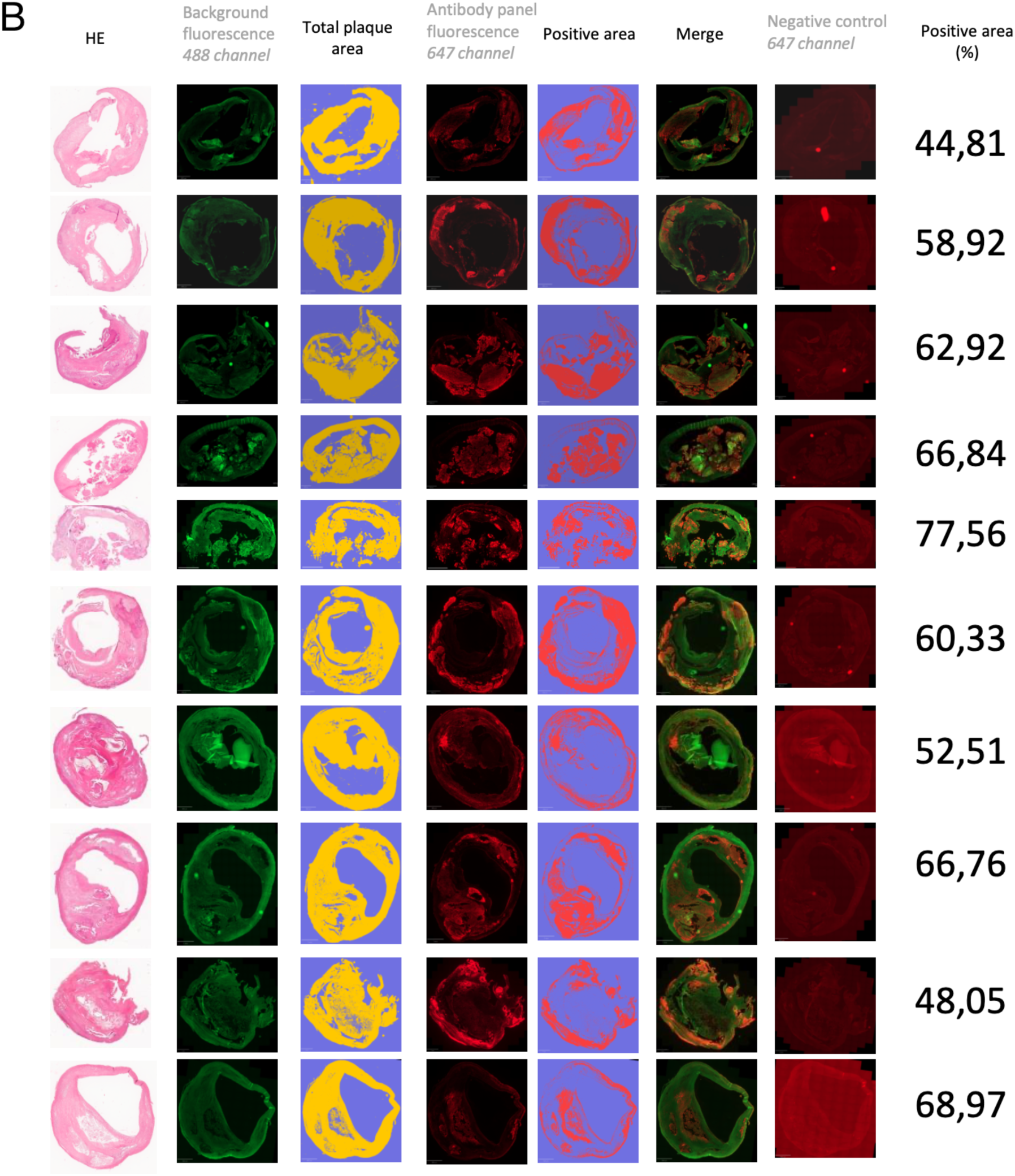
Individual immunofluorescence analyses of protein panel expression across atherosclerotic plaque samples. Representative images showing multiplexed immunofluorescence analysis of n=20 independent plaque samples. Stable plaques are shown in A, and vulnerable plaques are shown in B. For each sample, sequential columns show: H&E staining for tissue morphology, tissue autofluorescence (488 channel, green), computed total plaque area mask (yellow overlay on blue background), protein panel immunofluorescence (647 channel, red), segmented positive staining regions (red overlay on blue background), merged channels, and antibody negative control (647 channel). Positive area (%) indicates the proportion of total plaque area showing protein panel expression. Images demonstrate consistent protein panel detection across samples with minimal background signal in negative controls.

## Methods

### Ethics approval

Informed written consent for biobank sample contributions was obtained from all participants in accordance with the principles of the Declaration of Helsinki. The study protocol was approved by the local ethics committee of the Technical University of Munich (DigiMed ethics 2023-297; approval no. 2799/10). Clinical data were retrieved from anonymised electronic patient records.

### Sample collection and processing for biobank

#### Plaques tissue

Carotid plaques were obtained from patients undergoing carotid endarterectomy (CEA). Immediately after surgical retrieval, plaque samples were transferred into RNAlater (Thermo Scientific, Germany) to preserve RNA during transport and subsequent processing. Plaques were fixed in 4% paraformaldehyde (PFA) for 24 hours and decalcified with Entkalker soft SOLVAGREEN® (Carl ROTH, Karlsruhe, Germany) for 5 days. Following decalcification, plaque tissues were embedded in paraffin and stored as formalin-fixed paraffin-embedded (FFPE) blocks.

#### Serum samples

Blood was collected from CEA patients using S-Monovettes (Starstedt, Nümbrecht). All collected blood samples were allowed to clot at room temperature for 15 minutes. Subsequently, the clotted blood was centrifuged at 2000-rcf at 4 C for 10 minutes, and the supernatant serum was obtained and stored at -80 C for further processing.

### Cohort design

From 524 patients, 132 plaque samples and 505 serum samples were collected. Of these, 112 plaque samples and 505 serum samples were used for proteomics, while the remaining 20 were immunofluorescence validation samples. A subset of 93 patients provided plaque and serum samples for proteomic analyses (Table 1, Supplementary Table 1).

Plaque samples (n=112 for proteomics; n=20 for immunofluorescence) were selected based on the histological availability of the fibrous cap and medial layer, along with comprehensive clinical data. Serum samples (n=505) were selected based on sufficient volume, never previously thawed, low hemolysis and availability of complete clinical data. Case selection was optimised to maintain a covariate balance between stable and vulnerable patient subgroups.

### Histomorphology guided spatial proteomics of plaque tissues

#### Histological analysis

Formalin-fixed, paraffin-embedded (FFPE) plaque specimens were sectioned at 2 μm thickness and mounted on glass slides (Menzel SuperFrost, 76 × 26 × 1 mm, Fisher Scientific, Schwerte, Germany). Following deparaffinisation and rehydration, sections were stained using hematoxylin and eosin (HE) (ethanolic eosin Y solution and Mayer’s acidic hemalum solution, Waldeck, Münster, Germany) or Elastica van Gieson (EvG) (picrofuchsin solution [Romeis 16th edition] and Weigert’s solution I [Romeis 15th edition]) according to the manufacturer’s protocols. Slides were coverslipped with Pertex (Histolab Products, Askim, Sweden) and glass coverslips (24 × 50 mm, Engelbrecht, Edermünde, Germany). The H&E-stained slides were digitally scanned and used for subregion annotation.

A pathologist annotated the digital scans of the tissues to highlight three key subregions—media, fibrous cap, and necrotic core—using established morphological criteria. The media was identified as the arterial wall layer composed mainly of smooth muscle cells and elastic fibres. The necrotic core was characterised per AHA guidelines^1–4^and noted for its lipid-rich content, cellular debris, calcification, and areas of intraplaque haemorrhage. Fibrous cap thickness served as a measure of plaque vulnerability; caps measuring ≥200 μm were classified as stable, whereas those <200 μm were classified as unstable or vulnerable^5^.

#### Laser microdissection slide preparation

Two-micrometer PEN membrane slides (MicroDissect GmbH) were exposed to UV light at 254 nm for 1 hour and then coated with Vectabond (Vector Laboratories; SP-1800-7) according to the manufacturer’s protocol. Next, two consecutive 5-μm-thick FFPE sections from each block were mounted on the pretreated slides and dried overnight at 37 °C. In parallel, two additional 7-μm-thick sections were also placed on the same type of pretreated PEN membrane slides and dried under the same conditions.

After drying, slides were heated to 56 °C for 20 minutes to soften the paraffin. This was followed by deparaffinisation and dehydration using xylene (2 × 2 minutes) and a graded ethanol series (100%, 90%, 75%, and 50% ethanol, each for 2 × 1 minute). The slides were then air-dried briefly, ensuring the tissue was fully prepared for subsequent laser microdissection.

#### Laser microdissection

Digitally annotated H&E scans of each plaque were used as a reference and aligned with the corresponding FFPE sections on a Leica LMD7 laser microdissection microscope (Leica Microsystems, Germany). Within the annotated subregions, tissue areas with minimal intraplaque haemorrhage were prioritised for microdissection. For each subregion, 25 circles (area = 14,000 μm² each) were excised from two consecutive microtome sections and collected directly into the wells of a low-binding 96-well plate positioned beneath the slides. Laser cutting was performed with a power of 57, aperture 1–2, speed 23, and a middle-pulse setting. The plates were sealed with adhesive foil (Covaris), centrifuged at 1,000 × g for 2 minutes, and stored at −20 °C until further processing.

#### Sample preparation for mass spectrometry

All 336 samples were processed in a single batch to minimise potential batch effects. A semi-automated, high-throughput sample preparation method was employed using a Bravo pipetting robot (Agilent). First, each well was washed with 50 μL of 100% acetonitrile and dried in a SpeedVac at 25 °C for 30 minutes. Next, 40 μL of 60 mM triethylammonium bicarbonate (TEAB, Sigma) in MS-grade water was added to each well. The plates were then sealed with two layers of adhesive foil and heated at 95 °C for 60 minutes in a 96-well thermal cycler (Eppendorf).

After heating, 10 μL of 60% acetonitrile in 60 mM TEAB (resulting in a final acetonitrile concentration of 12.5%) was added, followed by heating at 75 °C for 60 minutes. The samples were then pre-digested with 40 ng LysC for 4 hours at 37 °C, followed by overnight digestion with 60 ng trypsin in the same 96-well thermal cycler. After approximately 16 hours, digestion was quenched by adding 15 μL of 6% trifluoroacetic acid (TFA) to achieve a final TFA concentration of 1%. Peptides were desalted using the iST 96x kit (PreOmics, Germany) according to the manufacturer’s instructions, dried in a SpeedVac, and stored at −20 °C until further analysis by mass spectrometry.

#### LC-MS/MS data acquisition

Dried peptide samples were reconstituted in 6 μL of MS loading buffer (2% acetonitrile [v/v], 0.1% trifluoroacetic acid [v/v] in MS-grade H₂O). Since plaque samples were stratified into subregions and distributed across four plates, a custom Python script was employed to track and randomise sample strata. The samples were transferred into new plates using a custom protocol on the OT-2 (OpenTrons) liquid handling system.

An EASY-nLC 1200 (Thermo Fisher Scientific) was coupled to a timsTOF Ultra mass spectrometer (Bruker, Germany) via a nanoelectrospray ion source (CaptiveSpray, Bruker). Peptides were separated on a 50 cm in-house–packed nano-HPLC column (75 μm inner diameter) containing 1.9 μm ReproSil-Pur C18-AQ silica beads (Dr. Maisch GmbH). The column was maintained at 60 °C in a custom-built column oven. A 120-minute linear gradient was delivered at a flow rate of 300 nL/min, starting from 3% to 30% buffer B over 95 minutes, increasing to 60% buffer B over 5 minutes, washing at 95% buffer B for 10 minutes, and re-equilibrating at 5% buffer B for 10 minutes. Buffer A consisted of 0.1% formic acid (FA) in 99.9% ddH₂O, whereas buffer B consisted of 0.1% FA, 80% acetonitrile (ACN), and 19.9% ddH₂O.

The timsTOF mass spectrometer was operated in dia-PASEF mode with DIA windows customised based on pilot experiments using plaque samples. The DIA windows covered an m/z range of 350–1200 and an ion mobility range of 0.7–1.3 Vs/cm. All other parameters were set to default as recommended^6^.

### Bioinformatic preparation of the proteomics data

#### LC-MS/MS data analysis

Peptide and protein identification was performed using a library-free search in DIA-NN, employing the UniProt FASTA database (Canonical, Version 2022-07-27, 20,420 sequences)^7^. Raw MS files from each plaque subregion were processed separately. Methionine oxidation was set as a variable modification, allowing up to one missed cleavage. The precursor charge range was specified from 2+ to 4+, with a precursor mass range of 350–1,200 m/z and peptide lengths of 7–35 amino acids. Both mass and MS1 accuracies were set to 15 based on prior estimations. Isotopologue detection and match-between-runs (MBR) were enabled, and the neural network classifier was configured for single-pass mode. The ‘--relaxed-prot-inf’ option enabled more conservative protein grouping, with protein inference performed directly from the FASTA file. Library generation was performed using “Smart profiling,” cross-run normalisation was set to “RT-dependent,” and the quantification strategy was “Robust LC (high precision).”

#### Refined protein quantitation and filtering

Peptide quantification data were used to re-quantify protein groups with at least five peptides. For these protein groups, pairwise correlation distances among its constituent peptides were calculated using a weighted Pearson correlation approach, chosen for its robustness against spurious correlations. The weight vector (w) was derived from a bivariate Gaussian kernel density estimate implemented in Python (scipy.stats.gaussian_kde).

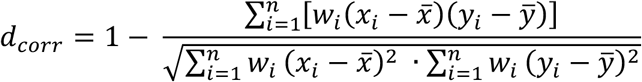

To identify a set of five highly correlated peptides per protein, agglomerative hierarchical clustering with average linkage was applied, followed by dynamic tree cutting within a distance range of 0.05–0.81. The peptides in each cluster were selected to represent the protein, and final protein intensities were recalculated using the MaxLFQ algorithm.

#### Protein and patient filtering and missing value imputation

Proteins with fewer than 27 observations and samples with fewer than 1,500 quantified proteins were excluded from further analysis. Missing values were imputed using a Python-based adaptation of the multiple imputation by chained equations approach^8^, implemented via IterativeImputer in scikit-learn. Several regression estimators were evaluated, including k-nearest neighbours, extra-trees, and random forests. Imputation performance was assessed by comparing the Frobenius norm of the correlation matrices derived from each imputed dataset against the unimputed reference matrix. Random forest-based imputation consistently outperformed other methods and was used for all subsequent analyses.

### Vulnerability prediction from the clinical metadata

To predict plaque vulnerability from clinical data, binary and nominal variables (e.g., sex, symptomatic status, diabetes) were one-hot encoded, whereas histological variables were treated as ordinal features. Continuous features (e.g., age, stenosis, BMI) were transformed into seven quantile bins. This feature transformation reduces the likelihood of overfitting and artificial splitting from high-cardinality variables.

For the plaque cohort (n=112), clinical metadata were randomly partitioned into a training set (75%) and a test set (25%). Model hyperparameter optimisation was performed via Bayesian optimisation on five-fold cross-validation of the training set. Three different algorithms were tuned: Random Forest (n_estimators, max_depth, min_samples_split, min_samples_leaf, max_features), XGBoost (colsample_bytree, eta, gamma, max_depth, min_child_weight, n_estimators), and HistGradientBoost (learning_rate, max_iter, max_leaf_nodes, max_depth, min_samples_leaf).

The optimised models were evaluated on the plaque test set (n=28) and an independent serum cohort (n=412), with performance measured as the area under the receiver operating characteristic (ROC) curve.

### Clustering of proteome data

#### Patient clusters

Recursive consensus clustering was performed on patients using the top 66% of most variable proteins^9^. A total of 100,000 iterations of consensus clustering were conducted with the following parameters: top_value_method=ATC, partition_method=skmeans, mean_silhouette_cutoff=0.9, min_samples=0.15, sample_by=row, and p_sampling=0.8. Adjusted mutual information (AMI) was used to assess the association between clinical covariates and the resulting plaque classifications. Statistical significance was evaluated via a permutation test (n=500,000) using AMI as the test statistic.

#### Protein clusters

Weighted gene correlation network analysis was performed on the full proteomic dataset to identify protein co-expression modules^10^. The power parameter (ranging from 1-20) for soft-thresholding was optimised to approximate a scale-free topology with a degree of independence of 0.85. A signed correlation was used in the adjacency matrix to capture both positive and negative correlations. The minimum module size was set to 50 proteins, and modules were merged if their distance height was less than 0.25.

#### Overrepresentation analysis

Enrichment of biological processes in the module genes was tested using g:Profiler^11^, using the following parameters: significant only, GO:BP, GO:CC, KEGG, REAC databases, and significance threshold set to FDR). P-value was determined using a hypergeometric test and was adjusted using Benjamini-Hochberg FDR. The seven most significant term names were selected and simplified into keywords using Sonnet 3.5 large language model (Anthropic, USA).

### Comparative statistical analysis

#### Binary comparisons

All binary comparative analysis (such comparison of subregions, stable vs. vulnerable, and POVPC and PCSK9 knockdown) were performed using the LIMMA package in R, called into python using.

#### Multiple Testing Correction

Statistical significance is reported with correction for multiple hypothesis testing. Nominal p values are reported as P. Statistical significance adjusted using the Benjamini–Hochberg procedure is denoted as FDR, whereas Q indicates p values adjusted using the Storey procedure^12^. Finally, FDR_IHW_ denotes p-value correction via independent hypothesis weighting, with the median intensity of each protein group serving as the covariate^13^.

### Enrichment map analysis

Pathways were defined using the gene set file Human_GOBP_AllPathways_no_GO_iea_June_03_2023_symbol.gmt^14^. Gene Set Enrichment Analysis (GSEA) used log2FC as the ranking metric, restricting the gene set size to 10 and 500^15^. A total of 5,000 permutations were performed. Enriched pathways were visualised using the EnrichmentMap app in Cytoscape^16^. Network maps were generated for nodes with FDR q < 0.01 and P < 0.0001; nodes sharing gene overlaps with a Jaccard coefficient > 0.25 were linked by a green edge. Clusters of related pathways were identified and annotated using a Markov Cluster algorithm (AutoAnnotate v1.2), which groups pathways based on shared keywords in the pathway description. The resulting pathway clusters are denoted as major pathways and displayed in a group.

### Vascular smooth muscle cell (VSMC) proteomes

Human carotid artery smooth muscle cells (HCtASMC) were purchased from PeloBiotech (PB-3514-05a, Lot: 3003, Planegg, Germany) and maintained in VSMC Medium (Smooth Muscle Cell Growth Medium, PB-MH-200-2100, PeloBiotech, Planegg, Germany) until further use.

#### Cell culture and treatment

Human vascular smooth muscle cells (VSMCs) were seeded at a density of 15,000 cells per well in 6-well plates using a standard VSMC medium. After 24 hours, the medium was discarded, and the cells were washed three times with phosphate-buffered saline (PBS), followed by three washes with serum-free, phenol red-free medium (SF&PRF; PB-MH-200-2190-FCS-PRF, PeloBiotech, Planegg, Germany). Cells were treated with 10 μg/mL POVPC or an equivalent volume of ethanol (solvent control) in SF&PRF medium for 72 hours.

At the end of the 72-hour treatment, the culture supernatant was collected (see harvest protocol). Subsequently, 25 nM Inclisiran (HY-132591, MedChemExpress) or 25 nM negative control siRNA (AM4614, Ambion) was introduced in SF&PRF medium using RNAiMAX (13778-150, Thermo Fisher). Following an additional 24-hour incubation, the supernatant and the cells were harvested and prepared for proteomic analysis.

#### Supernatant Collection

Cell culture supernatant was transferred into 2-mL microcentrifuge tubes and centrifuged at 300 × g for 10 minutes at 4 °C to remove non-adherent cells. Approximately 95% of the supernatant was transferred to a fresh 2-mL tube and centrifuged at 2000 × g for 20 minutes at 4 °C to remove debris. Finally, ∼95% of the clarified supernatant was transferred to another 2-mL tube and stored at −80 °C.

#### Cell Harvesting

After removing the medium, cells were scraped in 1 mL of ice-cold TBS and transferred to a microcentrifuge tube. The cell suspension was centrifuged at 300 × g for 10 minutes at 4 °C, and the supernatant was discarded. The cell pellet was then washed with ice-cold TBS and centrifuged. This washing step was repeated twice more for a total of three washes. After the final wash, the supernatant was carefully removed, and the pellet was stored at −80 °C with minimal residual liquid.

#### Processing of cellular and secretomes for LC-MS/MS

Cell pellets were processed using the iST kit (PreOmics, Germany) following the manufacturer’s instructions. Briefly, cells were resuspended in 100 μL of the “lyse” buffer and lysed using a BeatBox magnetic homogeniser (PreOmics, Germany) for 20 minutes at the standard setting. Lysates were denatured by heating at 95 °C for 10 minutes, followed by 5 minutes of sonication (10 cycles of 30 seconds on/off) to shear DNA. Proteins were digested overnight using a Trypsin/LysC mixture. Peptides were desalted with the kit’s cartridges, dried, and reconstituted in 0.1% formic acid in water. Secretome samples were thawed and concentrated to 100 μL using a 7 kDa MWCO spin filter. An equal volume of lyse buffer was added to each sample, and the same denaturation, digestion, and desalting steps described above were applied.

#### LC-MS/MS analysis

Samples were loaded onto Evotip Pure tips (Evosep) and separated using an Evosep One system employing the standardised Whisper 40-sample-per-day (40SPD) method. Peptides were passed through a 15 cm column (75 μm i.d.) packed with 1.7 μm C18 beads (IonOpticks), heated to 50 °C, and then introduced into a timsTOF mass spectrometer (Bruker). Data acquisition settings followed previously described protocols, along with protein filtering and imputation.

#### Knowledge graph analysis

The top 250 upregulated genes were used for the knowledge graph analysis^17^. Enrichment was performed against the GO:BP and the Human Gene Atlas database. Only the top 14 terms from each database were exported.

#### Pathway activation analysis

PROGENy analysis was used to assess changes in cellular signalling pathway activity in VSMCs under three conditions: control, POVPC treatment, and PCSK9 knockdown with POVPC^18^. Specifically, log₂ fold-change values (control vs control, POVPC vs control, and [PCSK9 knockdown + POVPC] vs POVPC) were derived from the proteomic data. The top 50 genes for each pathway model were used in the PROGENy inference. An ANOVA test was then applied to compare pathway activity distributions across the three experimental conditions.

### Protein Panel Development for Plaque Vulnerability Prediction

#### Feature selection

A multi-step pipeline was devised to identify an optimal protein panel for predicting plaque vulnerability using proteomic data from distinct plaque subregions (media, fibrous cap, necrotic core) and their combinations. First, proteomic profiles were stratified into training (70%) and testing (30%) sets, repeating this partitioning for each subregion independently and for all subregions combined. Potential features were initially selected by filtering for nominal p values (using the Limma framework) and consistent upregulation in vulnerable plaques, generating a “stats-filtered” set.

#### Protein panel development

To determine the optimal panel size, 5,000 random protein subsets for each candidate size (3–20 proteins) were evaluated using an XGBoost model, resulting in a total of 90,000 area under the receiver operating characteristic (AUC) evaluations. The panel size at which the AUC plateaued was deemed optimal; for plaque data, this inflexion point occurred at seven proteins.

A genetic algorithm then optimised the specific composition of these seven proteins. Each generation included 6,000 candidate solutions, of which the top 2,000 were selected for mating. Four elite solutions were carried over unchanged, while mutation and crossover probabilities were initially set to 0.3 and 0.1, respectively, and rose to 0.8 and 0.6 after five generations of stagnant fitness. Within each generation, five-fold cross-validation was performed using an Extra Trees Classifier, and optimisation ceased after 20 consecutive generations without improvements in accuracy.

#### Protein panel validation

Finally, the best-performing seven-protein panel was validated with an XGBoost classifier whose hyperparameters—colsample_bytree, eta, gamma, max_depth, min_child_weight, and n_estimators—were tuned via Bayesian optimisation on five-fold cross-validation of the training data.

### Quantitative immunofluorescence

#### Immunofluorescence (IF) Staining

Sections were mounted on SuperFrost Plus slides (Thermo Fisher Scientific, Waltham, MA, USA) pre-coated with 0.1% poly-L-lysine (Sigma-Aldrich, St. Louis, MO, USA). Antigen retrieval was performed by boiling the slides in a pressure cooker with 10 mM citrate buffer (pH 6.0), and endogenous peroxidase activity was blocked using 3% hydrogen peroxide. Sections were then blocked for 1 hour with blocking buffer (5% horse serum, 1% BSA, 0.5% Triton-X100) and incubated overnight with primary antibodies diluted in 5% horse serum. After primary incubation, appropriate secondary antibodies (table below) were applied for 1 hour under identical blocking conditions. Autofluorescence quenching and nuclear staining with DAPI were then performed. Primary antibodies were validated individually and combined for panel validation, and proper controls were included for each target. Images were acquired using a Zeiss Axioscan7 (Carl Zeiss Microscopy GmbH, Jena, Germany) and processed with ZEN 3.3 software. The table below describes the antibodies used:

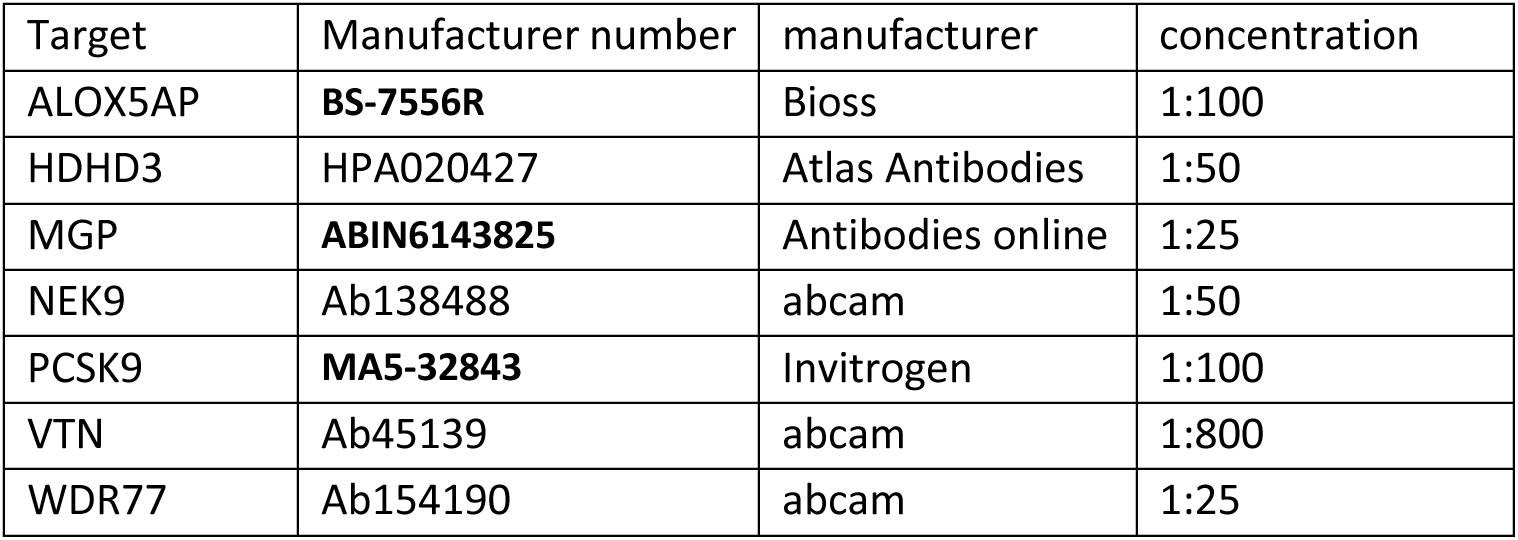

| Target | Manufacturer number | manufacturer | concentration |
| --- | --- | --- | --- |
| ALOX5AP | **BS-7556R** | Bioss | 1:100 |
| HDHD3 | HPA020427 | Atlas Antibodies | 1:50 |
| MGP | **ABIN6143825** | Antibodies online | 1:25 |
| NEK9 | Ab138488 | abcam | 1:50 |
| PCSK9 | **MA5-32843** | Invitrogen | 1:100 |
| VTN | Ab45139 | abcam | 1:800 |
| WDR77 | Ab154190 | abcam | 1:25 |

#### Quantitative analysis

Fluorescence images were analysed using QuPath^19^. First, autofluorescence in the 488 nm channel was inspected to delineate the entire plaque area. Next, a pixel classifier threshold was applied to the 647 nm channel to capture the antibody-specific signal from the biomarker panel, enabling accurate quantification of the stained region. All subsequent calculations and statistical analyses were performed using GraphPad Prism (v10, GraphPad Software, Boston, MA, USA).

### Serum proteome data

#### Sample preparation

Serum samples were processed with perchloric acid as described in the original publication^20^. Briefly, 5 μL of serum was diluted with 20 μL of water, followed by 25 μL of perchloric acid. The mixture was vigorously mixed for 1 hour at 4 °C, then centrifuged at 4,000 × *g* for 20 minutes at 4 °C. The supernatant was transferred to μSPE hydrophilic-lipophilic balance (HLB) plates (Waters, USA) and rebuffered into 50 mM ammonium bicarbonate. Proteins were digested into peptides using trypsin and Lys-C (1:100 enzyme-to-protein ratio). The resulting peptides were loaded onto Evotips for desalting and LC-MS/MS analysis. All liquid handling processing steps (aliquoting, pipetting, solution transfer, and rebuffering) were performed using Bravo liquid handling robot (Agilent, USA).

#### LC-MS/MS analysis

Peptides were separated on an EvoSep One system (Evosep) using the standard 60-samples-per-day (60SPD) method, which employs a 21-minute gradient. A 15 cm column (75 μm inner diameter) packed with 1.7 μm C18 beads (IonOpticks) was maintained at 50 °C and coupled to an Orbitrap Astral mass spectrometer (Thermo Fisher Scientific) via an EASY-Spray source. The spray voltage was set to 1,900 V. All samples were acquired in data-independent acquisition (DIA) mode at a resolution of 240,000, scanning from 380 to 980 m/z. The normalised AGC target was 500%, and an isolation window of 8 Th with a maximum injection time of 16 ms was used. Ions were fragmented at a higher-energy collisional dissociation (HCD) collision energy of 25%. Field asymmetric ion mobility spectrometry (FAIMS) was enabled at a compensation voltage of −40, and the gas flow was reduced to 2.5 L/min.

#### Data analysis

Data-independent acquisition (DIA) data were processed using DIA-NN as previously described. Peptides detected in fewer than 25% of samples were excluded, as were any samples with fewer than 6,000 quantified peptides. Protein groups were re-quantified using the correlation distance–based approach outlined earlier, and missing values were imputed using a random forest regressor in a multiple imputation framework.

### Serum biomarker panel development

A similar multi-step workflow was used to develop a serum biomarker panel, with a few modifications compared to the plaque-based approach. Serum samples were divided into two cohorts: matched (n=93; samples with corresponding plaque data) and independent (n=410). The matched cohort was used for initial feature selection, focusing on statistical significance, log₂ fold change (stable vs. vulnerable), and monotonic correlation of protein abundance between tissue and serum. The resulting set of proteins was then used to build a biomarker panel in the independent cohort, which was further split into training and testing sets.

The optimal panel size, determined to be 12 proteins, was identified via an information-content analysis similar to that performed for plaque tissues. Panel optimisation proceeded via a genetic algorithm, and the final model performance was validated in the test set.

